# A double index for assessing symptoms in psychosis based on acoustic and semantic information

**DOI:** 10.64898/2026.08.11.26360092

**Authors:** Maryia Kirdun, Rui He, Cemal Demirlek, José Tomás García-Molina, Roya Hüppi, Werner Surbeck, Noemi Dannecker, Burcu Verim, Berna Yalincetin, Victor Ortiz García de la Foz, Rosa Ayesa Arriola, Emre Bora, Alicia I. Figueroa-Barra, Filip Spaniel, Lena Palaniyappan, Iris E. Sommer, Philipp Homan, Wolfram Hinzen, Claudio Palominos

**Affiliations:** Department of Translation & Language Sciences, Universitat Pompeu Fabra, Barcelona, Spain; Department of Psychiatry, McLean Hospital, Harvard Medical School, Belmont, Massachusetts, USA; Department of Psychiatry and Mental Health, South Campus, School of Medicine, University of Chile, Santiago, Chile; Department of Adult Psychiatry and Psychotherapy, University of Zurich, Zurich, Switzerland; Department of Neurosciences, Health Sciences Institute, Dokuz Eylül University, Izmir, Türkiye; Valdecilla Research Institute (IDIVAL), Santander, Spain; Centro de Investigación Biomédica en Red de Salud Mental (CIBERSAM), Instituto de Salud Carlos III, Madrid, Spain; Department of Psychiatry, Dokuz Eylül University, Izmir, Türkiye; National Institute of Mental Health, Klecany, Czech Republic; Douglas Mental Health University Institute, Department of Psychiatry, McGill University, Montreal, Quebec, Canada; Department of Psychology, McGill University, Montreal, Quebec, Canada; Department of Psychiatry, Schulich School of Medicine and Dentistry, Western University, London, Ontario, Canada; Department of Neuroscience, University Medical Center Groningen, Antoni Deusinglaan 2, room 117 Groningen, Netherlands; Neuroscience Center Zurich, University of Zurich and ETH Zurich, Zurich, Switzerland; Institució Catalana de Recerca i Estudis Avançats (ICREA), Barcelona, Spain

**Keywords:** Schizophrenia, Large Language Models, Word Embeddings, Acoustic Signal, Semantic Space

## Abstract

Recent computational approaches to speech in psychosis generate large multidimensional feature spaces capturing semantic and acoustic aspects of language production. However, the clinical relevance of individual measures often becomes difficult to interpret due to redundancy, interaction effects, and high intercorrelation among features. Building upon previous work constructing a single composite index derived from semantic features based on language model embeddings, we here build a second acoustic index derived from speech acoustic features. Our aim was to evaluate the differential performance of both indices in conjunction in PANSS symptom prediction in psychosis in a cross-linguistic setting, including positive symptoms (P1, P2, P3), negative symptoms (N1, N4, N6), and general measures (G5, G9). The dataset comprised five languages and 221 patients with schizophrenia spectrum disorder (SSD). Both indices showed predictive power for individual PANSS scores, while also demonstrating clinically important complementarity: semantic indices were more strongly associated with positive symptom dimensions (P2, P3, Total Positive), whereas acoustic indices showed stronger relationships with negative and general symptoms (N1, N4, G5, G9). Both domains shared predictive overlap for global measures, such as PANSS Total scores. These findings suggest that both indices capture complementary and partially overlapping dimensions of psychopathology. The proposed composite index framework contributes to the advancement of low-dimensional speech-derived markers of symptom severity variation, potentially informing vulnerability to relapse and remission in psychosis.

## 1. Introduction

Alterations in speech and language, including disturbances in the semantic and grammatical organization as well as changes in acoustics and prosody, are among main manifestations in psychotic speech. Recent advances in computational methods now allow to automatically extract large multidimensional sets of semantic and acoustic features from clinical speech samples. However, this methodological richness also raises important challenges for interpretation, reproducibility and clinical translation, as the standard software and pipelines provide a larger number of features, which show strong correlation and some overlap. This generates a clear need for robust development of preprocessing pipelines that ensure clean, reliable and understandable use of the data.

Prior studies have focused on embedding-based measures of semantic properties derived from LLMs, typically operationalized through cosine similarity between word embeddings (Bedi et al., 2015; Bar et al., 2019; Pintos et al., 2022; Alonso-Sánchez et al., 2023; Palominos et al., 2024; He et al., 2024; Çokal et al., 2025). These studies have repeatedly reported altered semantic organization of speech in psychosis, often interpreted as reflecting a constrained semantic organization or atypical navigation in the semantic space (Nour et al., 2023; Palominos et al., 2024; He et al., 2024). Parallel work has studied acoustic properties of speech (De Boer et al., 2023; Teixeira et al., 2023; Ben Moshe et al., 2024; Worthington et al., 2025). Although semantic and acoustic approaches have largely developed separately, recent studies have begun to bridge these domains. For example, Voppel et al. (2023) showed that combining semantic and acoustic features improved classification of schizophrenia spectrum disorders (SSD) beyond either domain alone. Similarly, Rohanian et al. (2026) developed uncertainty-aware multimodal speech models integrating linguistic and acoustic information in a German-speaking sample, showing that multimodal fusion improved classification robustness, model calibration, and prediction of PANSS symptoms compared to unimodal approaches. Tang et al. (2025) examined 357 automated features spanning lexical, acoustic, and discourse, among others, in a longitudinal study, while Ciampelli et al. (2026) used a combination of acoustic and semantic measures to predict individual PANSS symptoms in a Dutch sample. Nevertheless, most work has emphasized diagnostic classification or group differences, relied on single-language datasets, and only rarely investigated symptom severity within SSD populations using integrated semantic and acoustic approaches.

One additional challenge is that the specific contribution of individual features within this high-dimensional space become difficult to interpret, as many measures may interact with one another or exhibit significant correlations. Rather than focusing on the isolated predictive value of individual features, an alternative approach is to model latent organizational properties that emerge from interactions among multiple features (Tang et al., 2025). Composite indices represent an attempt to exploit such higher-order structure by summarizing distributed patterns into lower-dimensional representations of the data (Nardo et al., 2005; Rockwood et al., 2007; Chakrabartty 2017; da Silva et al., 2026).

The present study expands upon previous work (Palominos et al., 2025), in which a composite semantic index showed its ability to distinguish patients with SSD from healthy controls and to longitudinally identify remission. Here, we further develop and evaluate this framework by constructing both semantic and acoustic indices and incorporating one additional information-theoretic measure of language predictability.

This allows us to compare their predictive and differential power, as well as examine their association with clinical symptoms. In contrast to the previous study (Palominos et al., 2025), which focused primarily on group differences using semantic indices derived from English-language data and tracked patients longitudinally, the current work centers on comparisons within the SSD group itself, with particular emphasis on the relationship between both acoustic and semantic indices and symptom severity. Moreover, the present study extends the analysis across five languages, providing a cross-linguistic perspective. Our general hypothesis was that indices may function as low-dimensional markers of latent psychopathological organization.

Our double index approach was specifically designed to enable tracking psychosis-related symptom variation and potentially help identify vulnerability to relapse or likelihood of remission. To this end, we targeted symptom dimensions specifically relevant to relapse prediction (as defined by Siafis et al., 2024), while the process of semantic and acoustic feature selection was guided by the goal of incorporating a comprehensive set of variables spanning the feature space commonly used in current computational analyses of speech in psychosis.

Previous studies have shown that acoustic measures can also be predictive of clinical outcomes (He et al., 2023; He et al., 2026), although not always specific to psychosis-specific outcomes such as PANSS scores, and frequently studied in broader diagnostic or classification contexts (Haider et al., 2020; Agurto et al., 2022; Pokorny et al., 2022; Berardi et al., 2023; García-Gutiérrez et al., 2023). By jointly studying both indices, we seek to determine whether these two domains provide complementary information regarding psychopathology and symptom expression. These proposed indices are not intended to map directly onto individual symptoms in a one-to-one manner. Instead, they are designed to capture broader latent patterns in speech organization that together may reflect underlying dimensions of psychopathology.

## 2. Methods

### 2.1 Participants

Both inpatients and outpatients were recruited in different studies from five specialized psychiatric centers in the Czech Republic, Spain, Chile (Figueroa-Barra et al., 2022), Switzerland (Surbeck et al., 2025; Rohanian et al., 2026; Hüppi et al., 2026), and Türkiye (Arslan et al., 2024), with approval from each site’s respective ethics committee. All cohorts included individuals diagnosed with schizophrenia spectrum or related psychotic disorders according to ICD-10 or DSM-IV criteria, including schizophrenia, schizo-affective disorder, brief psychotic disorder and psychosis not otherwise specified. Among common exclusion criteria across the sites were neurological disorders, significant medical conditions, intellectual disability, substance dependence or current substance abuse, and other factors potentially affecting neurocognitive or language assessments. Detailed inclusion and exclusion criteria for each site are provided in **Supplementary Table S1**.

The final compiled multilinguistic dataset comprised 221 participants across five languages: Czech (n = 16), Spanish (n = 48), Chilean-Spanish (n = 14), Swiss-German (n = 35), and Turkish (n = 108). Mean age ranged from 19.36 years (SD = 2.79) in the youngest Chilean Spanish cohort to 39.75 years (SD = 9.43) in the Spanish cohort. The proportion of female participants varied between 25% and 50% across datasets, the Spanish cohort being the most balanced. Years of education ranged from 11.0 (SD = 3.51) in the Spanish sample to 14.75 (SD = 3.62) in the Czech sample; education data were not available for Swiss German.

Diagnoses were determined by trained local psychiatrists based on structured diagnostic interviews. Symptom severity was assessed using PANSS (Positive and Negative Syndrome Scale) assessments, except for Turkish where SAPS scale (Scale for the Assessment of Positive Symptoms) was applied. SAPS scores were converted to positive PANSS scores using the method of Grot et al. (2021) for comparison with other cohorts. We focused on eight PANSS items available in all cohorts except Turkish: three positive (P1 – Delusions, P2 – Conceptual Disorganization, P3 – Hallucinatory Behavior), three negative (N1 – Blunted Affect, N4 – Passive/Apathetic Social Withdrawal, N6 – Lack of Spontaneity and Flow of Conversation), and two general items (G5 – Mannerisms and Posturing, G9 – Unusual Thought Content). Only positive items were available for the Turkish cohort (P1, P2, and P3). These eight items were selected as they form the core of the RSWG (Remission in Schizophrenia Working Group) PANSS score-based definition of symptomatic remission in schizophrenia (consensus agreement by Siafis et al., 2024). The total score for each subscale was calculated by summing the selected individual items, and the overall PANSS total was then calculated by summing the three subscale totals. The Chilean Spanish cohort showed the highest overall symptom levels (PANSS Total = 25.07, SD = 2.7); whereas the Spanish chronic cohort showed the lowest total scores (PANSS Total = 11.52, SD = 4.82), reflecting the greater clinical stability expected in a chronic sample. Demographic and clinical information for all the participants is provided in **Table 1**.

**Table 1:**
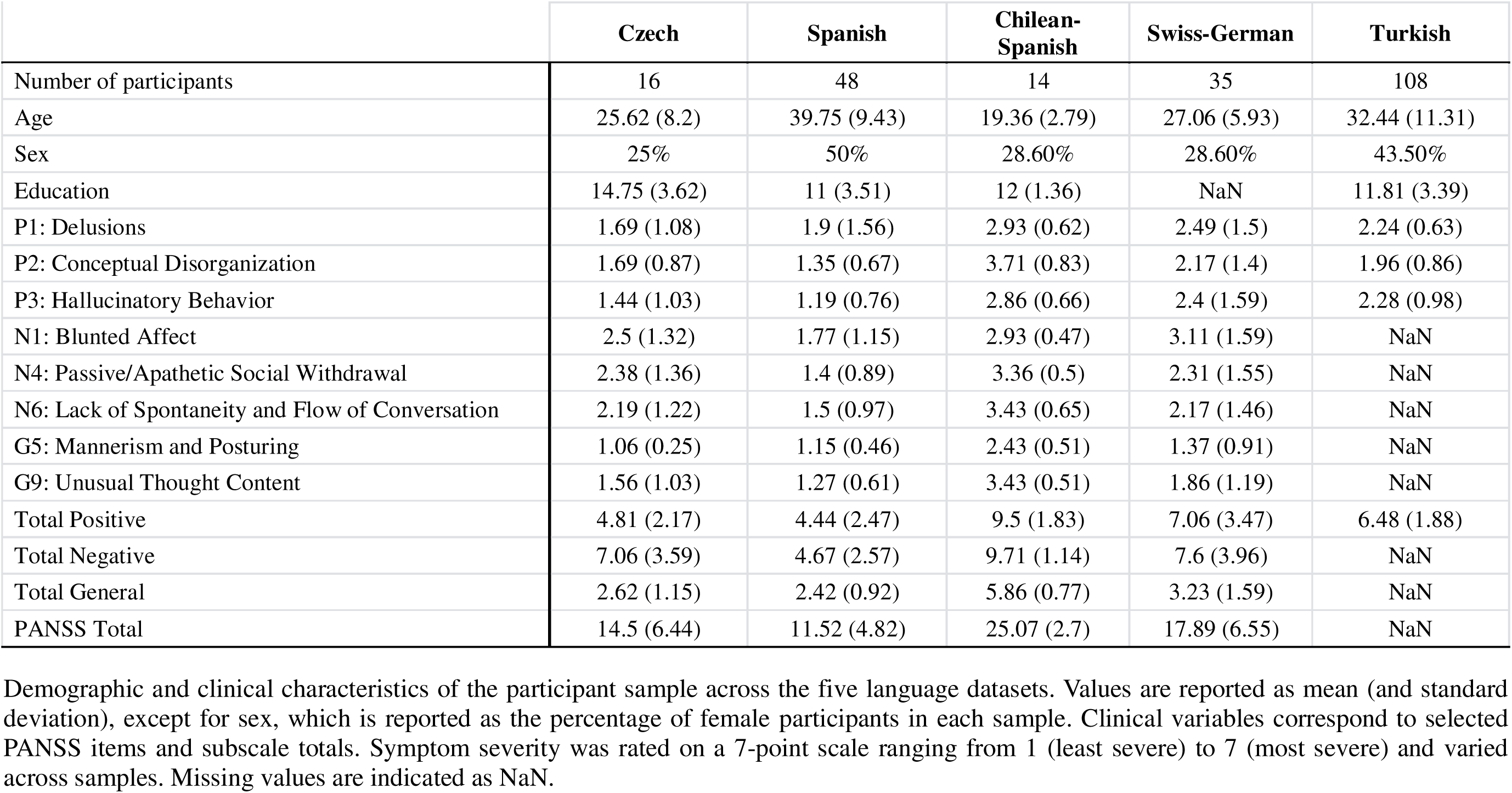
Demographic and clinical characteristics of the participants.

### 2.2 Data collection and preprocessing

Participants across all sites were audio-recorded while completing speech tasks. The Czech, Spanish, Turkish and part of the Swiss-German cohorts followed Discourse in Psychosis Speech Bank Protocol (https://discourseinpsychosis.org/), described in Melshin et al. (2025), which enables a consistent cross-linguistic investigation of language patterns in psychosis. The protocol includes free speech, picture descriptions, cartoon storyboard, dream narration, and a story reading and recall. The Chilean-Spanish participants completed clinical interviews and a subset of the Swiss-German cohort had to describe fourteen different pictures. Recordings were split by tasks and transcribed by human annotators.

Speech transcripts were pre-processed in Python (version 3.13) to extract the speaker’s speech only, and removing other information. Language-specific NLP pipelines were implemented by using Stanza for Czech and Turkish, and spaCy for Spanish, Chilean-Spanish and Swiss-German. The transcripts were tokenized at two levels: all tokens and content words only, restricted to nouns, verbs, adjectives, and adverbs.

Semantic and acoustic data were initially matched on observations, retaining only tasks with both transcripts and usable audio recordings. Observations with missing transcripts, corrupted or low-quality audio, or missing clinical scores were excluded. A total of 1513 semantic (Czech = 115, Spanish = 190, Chilean-Spanish = 95, Swiss-German = 350, Turkish = 763) and 1486 acoustic observations was finally included (Czech = 115, Spanish = 190, Chilean-Spanish = 95, Swiss-German = 323, Turkish = 763). The discrepancy in the number of Swiss-German observations relative to the other languages reflect differences in task administration: in the Swiss-German dataset, all picture descriptions were recorded as a single audio file and later split into multiple transcripts.

### 2.3 Feature extraction

#### 2.3.1 Semantic measures

Pre-trained models were employed for automated language processing. For word-level analyses, we used static fastText embeddings (Grave et al., 2018) and contextual BERT (Bidirectional Encoder Representations from Transformers; Devlin et al., 2019). FastText provides flexible static embeddings, effectively handling out-of-vocabulary and rare word forms through subword modelling (Bojanowski et al., 2017); whereas BERT generates bidirectional contextualized embeddings by considering both preceding and following words, allowing to capture richer contextual information. Language-specific BERT models were applied for Spanish, Chilean-Spanish, Swiss-German and Turkish; whereas a multilingual BERT model was used for Czech due to the absence of a language-specific model. For sentence-level representations, we used a multilingual SBERT (Sentence-BERT) model (Reimers & Gurevych, 2019). Finally, LLaMa 3.1 8B multilingual model (Meta AI, 2024) was used to calculate perplexity scores. Using these four models helps to cover different kinds of semantic meaning: lexical similarity, local contextual coherence, sentence and discourse-level coherence.

We adapted the semantic feature calculation pipeline from previous work (Palominos et al., 2025) and introduced perplexity as a new information-theoretic measure that quantifies predictability of a sequence of words according to a language model. First, we extracted fastText, BERT and SBERT embeddings on the word and sentence levels for each participant and task. Only content words were embedded in fastText. BERT embeddings were computed from full sentences including punctuation and stop words to preserve contextual and syntactic information, after which special tokens and punctuation were removed for semantic similarity calculation. Sentence embeddings were calculated in SBERT over the sentences. After generating the embeddings, cosine semantic similarities were computed between content words, between all words, and between sentences. To avoid bias from repeated words or sentences, fastText- and SBERT-based similarity values equal to 1 were excluded. Dynamic and static semantic features were derived from computed semantic similarities, and perplexity was calculated over the whole text. In total, 118 semantic features were extracted. A complete description of all semantic features is provided in **Table 2**.

**Table 2.**
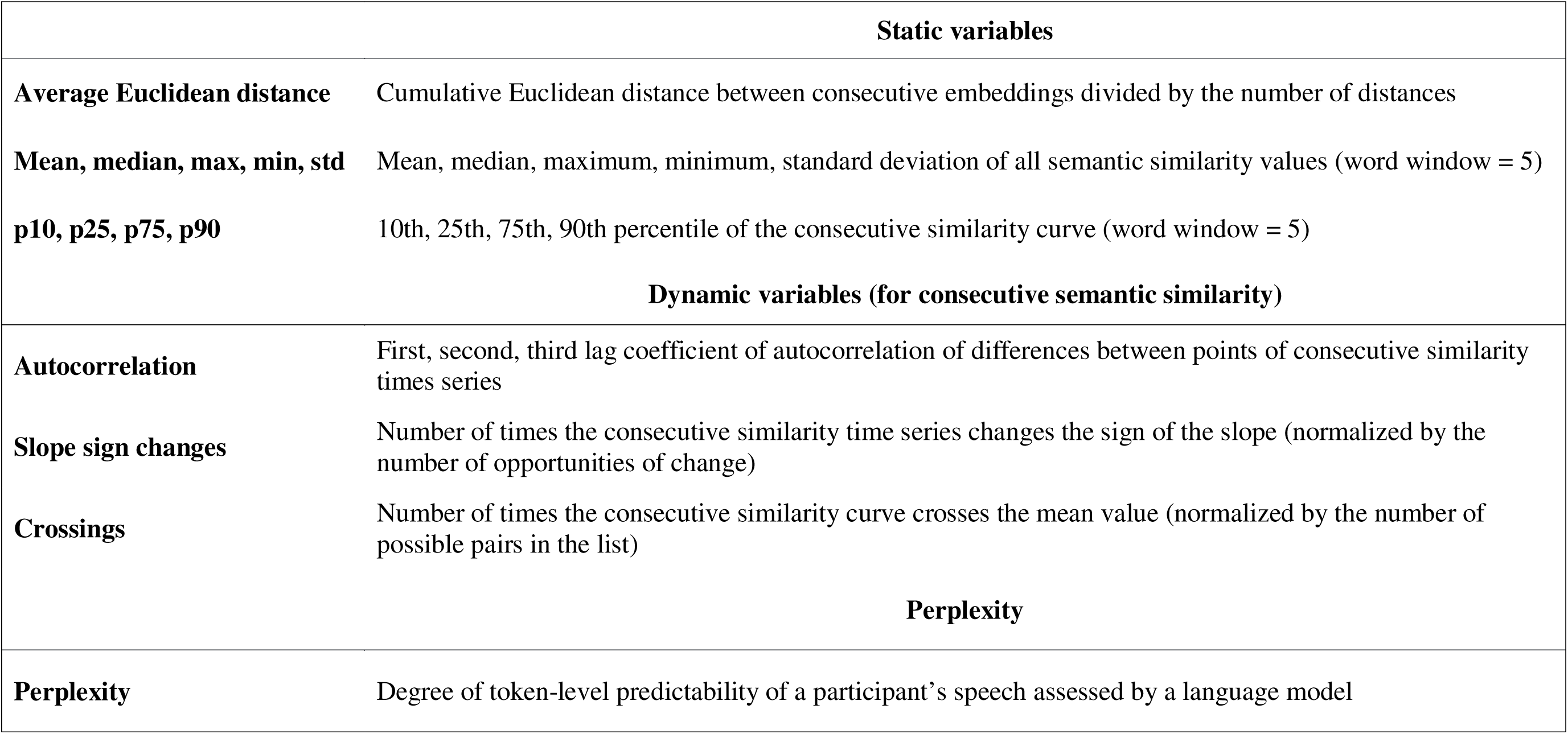
Semantic features calculated to build the semantic composite index.

#### 2.3.2 Acoustic measures

Acoustic features were extracted using the extended Geneva Minimalistic Acoustic Parameter Set (eGeMAPS) introduced by Eyben et al. (2016) within the openSMILE toolkit (Eyben et al., 2010; Eyben et al., 2013). This standardized limited feature set comprises 88 features capturing frequency, energy, spectral and temporal characteristics at the utterance level (24 frequency-related, 43 spectral/cepstral, 15 energy/amplitude-related, and 6 temporal features). The eGeMAPS are widely applied in automatic voice analysis, including paralinguistic and clinical speech research due to their suitability for automatic extraction, ease in calculation, demonstrated theoretical relevance, and sensitivity to affective and physiological changes in speech production (Pokorny et al., 2022; Xue et al., 2019). The list of all 88 acoustic features is provided in **Supplementary Table S2**.

### 2.4 Principal Component Analysis and composite index calculation

Similar to Palominos et al. (2025), the current workflow focused on constructing composite indices, here extended to both semantic and acoustic domains through feature selection and dimensionality reduction procedures. The pipeline combined preprocessing, redundancy reduction, principal component analysis (PCA), and the construction of final composite indices summarizing speech characteristics across participants and tasks.

For the semantic features, missing values were first imputed separately within each language using the Iterative Imputer with a linear regression estimator from the *sklearn.impute* module in Python. In total, there were 553 missing values across 1513 observations and 118 variables. No imputation procedure was required for the acoustic features, as the dataset contained no missing values.

First, to avoid redundancy, across languages, highly correlated pairs of features were removed based on variance (80% threshold), with the lower-variance feature being dropped. After this step, the number of semantic features decreased from 118 to 64, and acoustic features from 88 to 57. PCA was then applied for dimensionality reduction. The number of retained components was determined through parallel analysis (Horn, 1965; Longman et al., 1989) with a 95% threshold. This resulted in 14 components for the semantic dataset (explaining 67% of the variance) and 10 components for the acoustic dataset (explaining 61% of the variance). The dimensionality reduction aimed to obtain two single composite indices (Nardo et al., 2005) respectively, summarizing the semantic or acoustic characteristics of each participant and task. These indices were computed as the weighted average of the retained principal components (Chao, 2017). Finally, the indices were z-score normalized and scaled to a 0–1 range for interpretability.

### 2.5 Statistical analysis

Statistical modelling was done in R version 4.5.2 (R Core Team, 2025). To initially assess correlations with PANSS scores, we computed a participant-level geometric mean of task indices. After that we ran two sets of models: one using the geometric mean indices and another using the original task-level indices.

First, to examine the association between the indices and symptom severity, we fitted a linear regression model predicting each PANSS score, using the log-transformed participant-level geometric mean index as the main predictor. The geometric mean was used to aggregate index values across different tasks because it is less sensitive to extreme values and better captures the central tendency. Age and sex were included as covariates to control for demographic effects. This model allowed us to assess whether participants with higher index values exhibit higher or lower PANSS scores after adjustment for demographic factors. The model specification is provided in Equation 1:

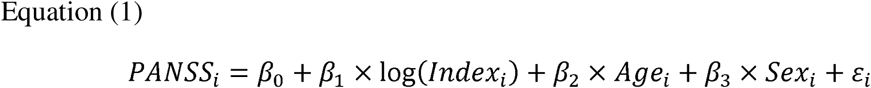

Secondly, we investigated how the index varies as a function of clinical symptoms and task conditions while accounting for repeated measurements. To this end, we fitted a generalized linear mixed-effects model (GLMM) with a beta distribution, as the index is bounded between 0 and 1. In this model, the dependent variable is the task-level index (not averaged across tasks), with repeated observations within participants. Symptom scores, task, age, and sex were included as fixed effects, and participant was modelled as a random intercept. The full model specification is provided in Equation 2:

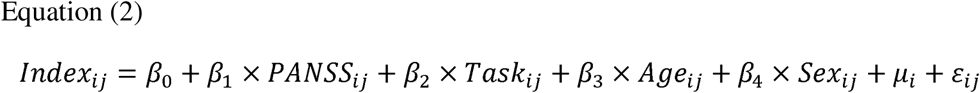

Finally, after obtaining the results for Equation (2), we examined which PANSS scores were significantly associated with both indices and which were specific to each index. To further characterize the relative contribution of each index for the PANSS scores shared by both indices, we performed a commonality analysis using participant-level geometric mean indices. Specifically, we modelled each PANSS score as a function of both the semantic and acoustic indices, while controlling for age and sex. The model specification is provided in Equation (3). The commonality analysis provides a way to decompose the incremental explained variance attributable to the indices into unique and shared components. This allowed us to estimate the proportion of variance uniquely explained by each index, and the variance jointly explained by both of them due to their correlation.

Additionally, we computed a correlation between both indices to quantify the degree to which they capture related aspects. We expected a positive but moderate correlation, rather than near-identity association, consistent with the hypothesis that both indices capture partially overlapping and complementary dimensions.

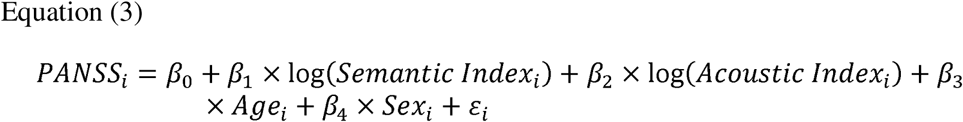

It is worth noting that, across all levels of analysis, the direction of the associations between PANSS scores and the indices, reflected in the correlation coefficients and the estimated values, is inherently arbitrary. This is because each index was constructed independently through dimensionality reduction, for which the resulting component sign has no intrinsic meaning. In practice, the sign of an index can be inverted without altering its statistical properties or interpretation. For this reason, the raw sign of the association should not be interpreted in absolute terms. At the same time, this does not mean that directionality is irrelevant for clinical interpretation. If the indices are to be used as tracking markers, their polarity must be fixed explicitly so the increases and decreases can be interpreted consistently.

## 3. Results

### 3.1 Correlations between PANSS scores and indices

A first analysis of the correlations between PANSS scores and the semantic and acoustic indices is presented in **Table 3**. We observed significant mild positive correlations between the semantic index and the P2, P3, N1, and G9 PANSS items. Also, the acoustic index showed significant mild to moderate correlations with the P2, N1, N4, N6, G5, and G9 items.

**Table 3:**
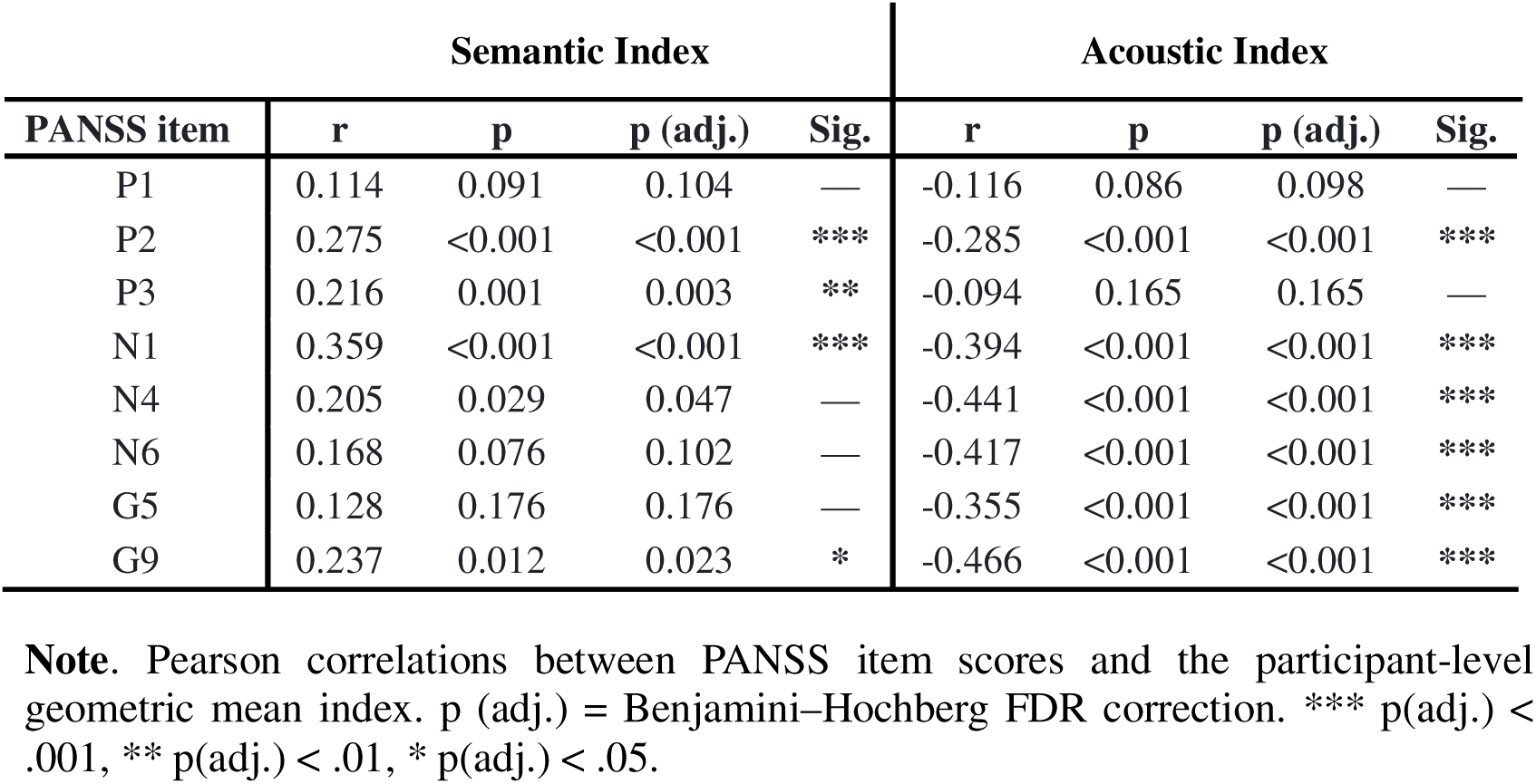
Correlations between PANSS symptom scores and semantic and acoustic indices.

| PANSS item | Semantic Index |  |  |  | Acoustic Index |  |  |  |
| --- | --- | --- | --- | --- | --- | --- | --- | --- |
|  | r | p | p (adj.) | Sig. | r | p | p (adj.) | Sig. |
| P1 | 0.114 | 0.091 | 0.104 | — | -0.116 | 0.086 | 0.098 | — |
| P2 | 0.275 | <0.001 | <0.001 | *** | -0.285 | <0.001 | <0.001 | *** |
| P3 | 0.216 | 0.001 | 0.003 | ** | -0.094 | 0.165 | 0.165 | — |
| N1 | 0.359 | <0.001 | <0.001 | *** | -0.394 | <0.001 | <0.001 | *** |
| N4 | 0.205 | 0.029 | 0.047 | — | -0.441 | <0.001 | <0.001 | *** |
| N6 | 0.168 | 0.076 | 0.102 | — | -0.417 | <0.001 | <0.001 | *** |
| G5 | 0.128 | 0.176 | 0.176 | — | -0.355 | <0.001 | <0.001 | *** |
| G9 | 0.237 | 0.012 | 0.023 | * | -0.466 | <0.001 | <0.001 | *** |

**Note.** Pearson correlations between PANSS item scores and the participant-level geometric mean index. p (adj.) = Benjamini–Hochberg FDR correction. \*\*\* p(adj.) < .001, \*\* p(adj.) < .01, \* p(adj.) < .05.

### 3.2 Linear regression analysis of PANSS scores and indices

**Table 4** shows the results of Equation (1), specifically the estimated coefficient, corresponding to the effect of the log-transformed geometric mean index on each PANSS score, for both the semantic and acoustic indices. For each regression model, we also report the statistical significance of. Full regression results, including all estimated coefficients and covariates, are provided in **Supplementary Table S3**.

**Table 4:**
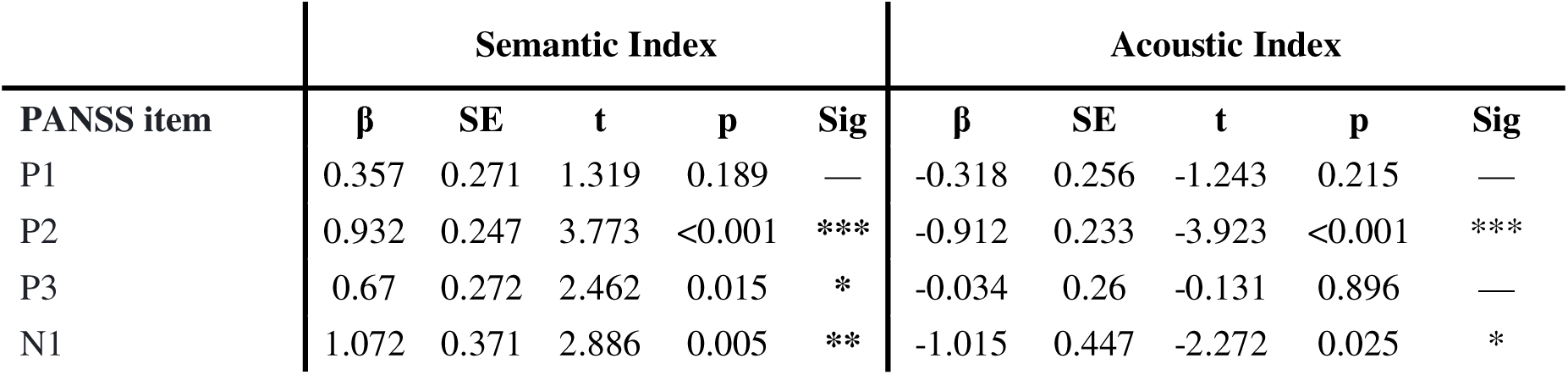
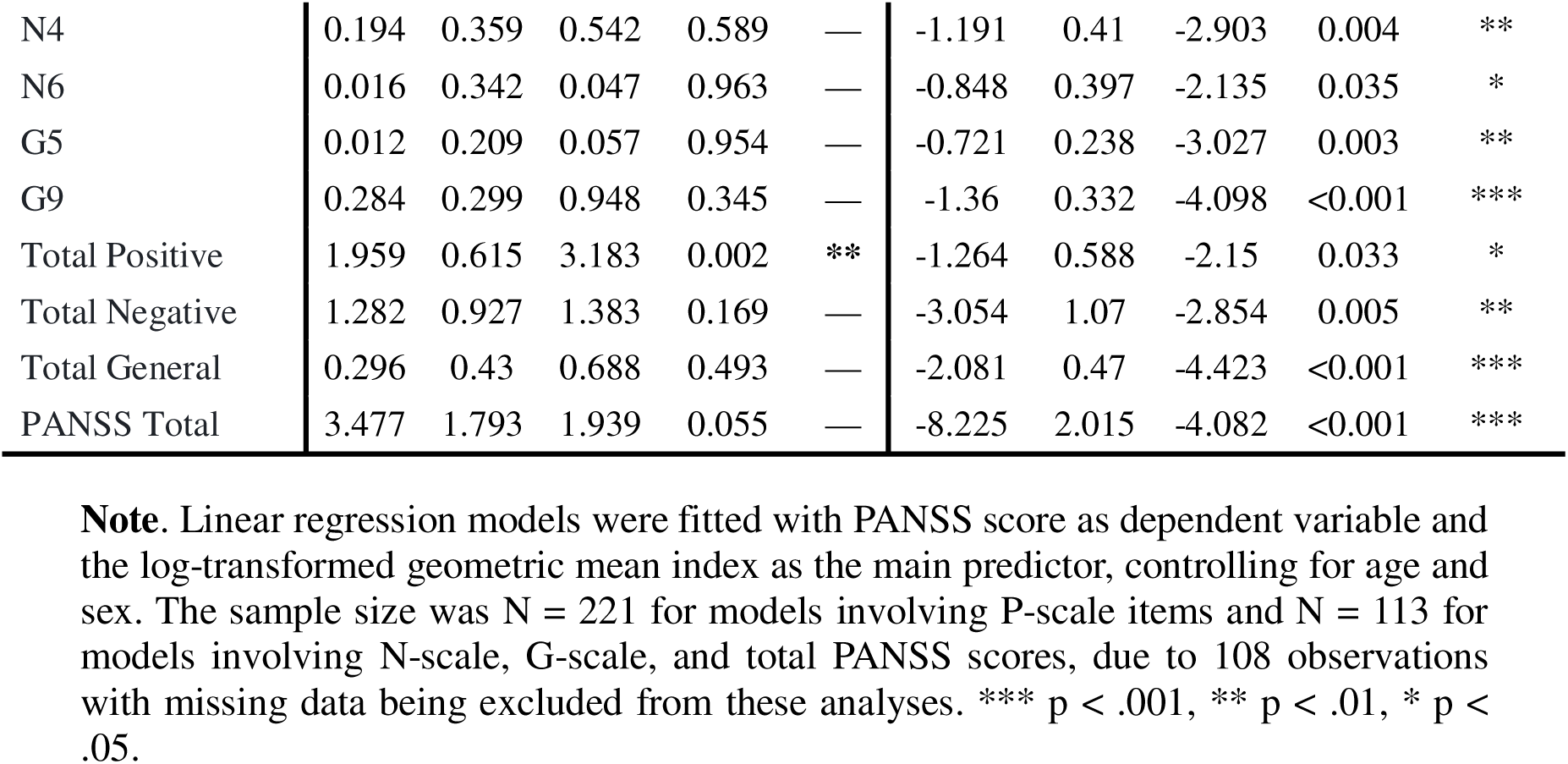
Linear regression results predicting PANSS scores using semantic and acoustic indices.

| PANSS item | Semantic Index |  |  |  |  | Acoustic Index |  |  |  |  |
| --- | --- | --- | --- | --- | --- | --- | --- | --- | --- | --- |
| | $\beta$ | SE | t | p | Sig | $\beta$ | SE | t | p | Sig |
| P1 | 0.357 | 0.271 | 1.319 | 0.189 | — | -0.318 | 0.256 | -1.243 | 0.215 | — |
| P2 | 0.932 | 0.247 | 3.773 | <0.001 | *** | -0.912 | 0.233 | -3.923 | <0.001 | *** |
| P3 | 0.67 | 0.272 | 2.462 | 0.015 | * | -0.034 | 0.26 | -0.131 | 0.896 | — |
| N1 | 1.072 | 0.371 | 2.886 | 0.005 | ** | -1.015 | 0.447 | -2.272 | 0.025 | * |
| N4 | 0.194 | 0.359 | 0.542 | 0.589 | — | -1.191 | 0.41 | -2.903 | 0.004 | ** |
| N6 | 0.016 | 0.342 | 0.047 | 0.963 | — | -0.848 | 0.397 | -2.135 | 0.035 | * |
| G5 | 0.012 | 0.209 | 0.057 | 0.954 | — | -0.721 | 0.238 | -3.027 | 0.003 | ** |
| G9 | 0.284 | 0.299 | 0.948 | 0.345 | — | -1.36 | 0.332 | -4.098 | <0.001 | *** |
| Total Positive | 1.959 | 0.615 | 3.183 | 0.002 | ** | -1.264 | 0.588 | -2.15 | 0.033 | * |
| Total Negative | 1.282 | 0.927 | 1.383 | 0.169 | — | -3.054 | 1.07 | -2.854 | 0.005 | ** |
| Total General | 0.296 | 0.43 | 0.688 | 0.493 | — | -2.081 | 0.47 | -4.423 | <0.001 | *** |
| PANSS Total | 3.477 | 1.793 | 1.939 | 0.055 | — | -8.225 | 2.015 | -4.082 | <0.001 | *** |

**Note.** Linear regression models were fitted with PANSS score as dependent variable and the log-transformed geometric mean index as the main predictor, controlling for age and sex. The sample size was N = 221 for models involving P-scale items and N = 113 for models involving N-scale, G-scale, and total PANSS scores, due to 108 observations with missing data being excluded from these analyses. \*\*\* p < .001, \*\* p < .01, \* p < .05.

For the semantic index models, age showed significant effects on PANSS scores in all models except those predicting the P1, P2, and N1 items. Notably, the effect of age was small and in the opposite direction to that of the semantic index. In the acoustic index models, age showed significant effects on PANSS scores, in the models predicting the N6 item and the Total Positive scores only, with small effects in the same direction as the acoustic index.

### 3.3 Task-level mixed-effects models predicting semantic and acoustic indices from PANSS scores

To account for repeated measures and differences across tasks, we fitted mixed-effects models predicting each index from PANSS scores (one model for PANSS score). Symptom scores, task, age, and sex were included as fixed effects, while participant was modelled as a random intercept. This allowed us to assess variation of indices while controlling for task-related effects and within-subject dependencies. The results of these models are presented in **Table 5**, while the full regression results, including all estimated coefficients and covariates, are provided in **Supplementary Table S4**, where task effects can be examined in detail.

**Table 5:** Mixed-effects results predicting indices scores from task-level semantic and acoustic indices.

| PANSS item | Semantic Index |  |  |  |  | Acoustic Index |  |  |  |  |
| --- | --- | --- | --- | --- | --- | --- | --- | --- | --- | --- |
| | $\beta$ | SE | z | p | Sig | $\beta$ | SE | z | p | Sig |
| P1 | 0.027 | 0.021 | 1.241 | 0.215 | — | -0.035 | 0.04 | -0.86 | 0.391 | — |
| P2 | 0.078 | 0.022 | 3.493 | <0.001 | *** | -0.122 | 0.042 | -2.87 | 0.004 | ** |
| P3 | 0.044 | 0.021 | 2.111 | 0.035 | * | 0.008 | 0.04 | 0.19 | 0.847 | — |
| N1 | 0.088 | 0.029 | 3.016 | 0.003 | ** | -0.091 | 0.035 | -2.56 | 0.011 | * |
| N4 | 0.018 | 0.033 | 0.539 | 0.59 | — | -0.095 | 0.038 | -2.49 | 0.013 | * |
| N6 | 0 | 0.035 | -0.011 | 0.991 | — | -0.067 | 0.041 | -1.65 | 0.1 | — |
| G5 | 0.014 | 0.057 | 0.239 | 0.811 | — | -0.149 | 0.066 | -2.25 | 0.025 | * |
| G9 | 0.047 | 0.039 | 1.201 | 0.23 | — | -0.142 | 0.045 | -3.14 | 0.002 | ** |
| Total Positive | 0.026 | 0.009 | 2.888 | 0.004 | ** | -0.025 | 0.017 | -1.43 | 0.152 | — |
| Total Negative | 0.018 | 0.012 | 1.42 | 0.156 | — | -0.038 | 0.015 | -2.62 | 0.009 | ** |
| Total General | 0.027 | 0.028 | 0.962 | 0.336 | — | -0.104 | 0.031 | -3.33 | <0.001 | *** |
| PANSS Total | 0.014 | 0.006 | 2.23 | 0.026 | * | -0.025 | 0.007 | -3.38 | <0.001 | *** |

**Note.** Mixed-effect models were fitted with the task-level index as the dependent variable and PANSS score as the main predictor, controlling for age, sex, and task (reference level = dream telling). Models were estimated using mixed-effects beta regression with a logit link and participant-specific random intercepts. Because beta regression requires values to lie strictly between 0 and 1, two observations with index values equal to 0 or 1 were excluded from the analyses. For the semantic index models, the sample size was N = 1513 observations across 221 participants for P-scale models, and N = 748 observations across 113 participants for N-scale, G-scale, and total PANSS models. For the acoustic index models, the sample size was N = 1486 observations across 221 participants for P-scale models, and N = 722 observations across 113 participants for N-scale, G-scale, and total PANSS models. Blue coefficients ( $\beta$ ) indicate positive associations between PANSS scores and index values, whereas red coefficients indicate negative associations. \*\*\* p < .001, \*\* p < .01, \* p < .05.

Notably, task effects were observed for all positive symptoms predictions when using the semantic index as the dependent variable, whereas no task effects were found for negative symptoms. In contrast, when using the acoustic index, task effects were limited to the picture description task for positive symptoms, and to the free speech task for negative and general symptoms. As noted previously, the resulting β coefficient signs are arbitrary.

As a summary of the results presented in **Table 5**, **Figure 1** shows a Venn diagram illustrating the PANSS scores significantly associated with the semantic and acoustic indices. The diagram highlights both the shared associations between the two indices and the symptom dimensions that were uniquely related to either semantic or acoustic features, providing an overview of their common and differential clinical correlates.

**Figure 1:**
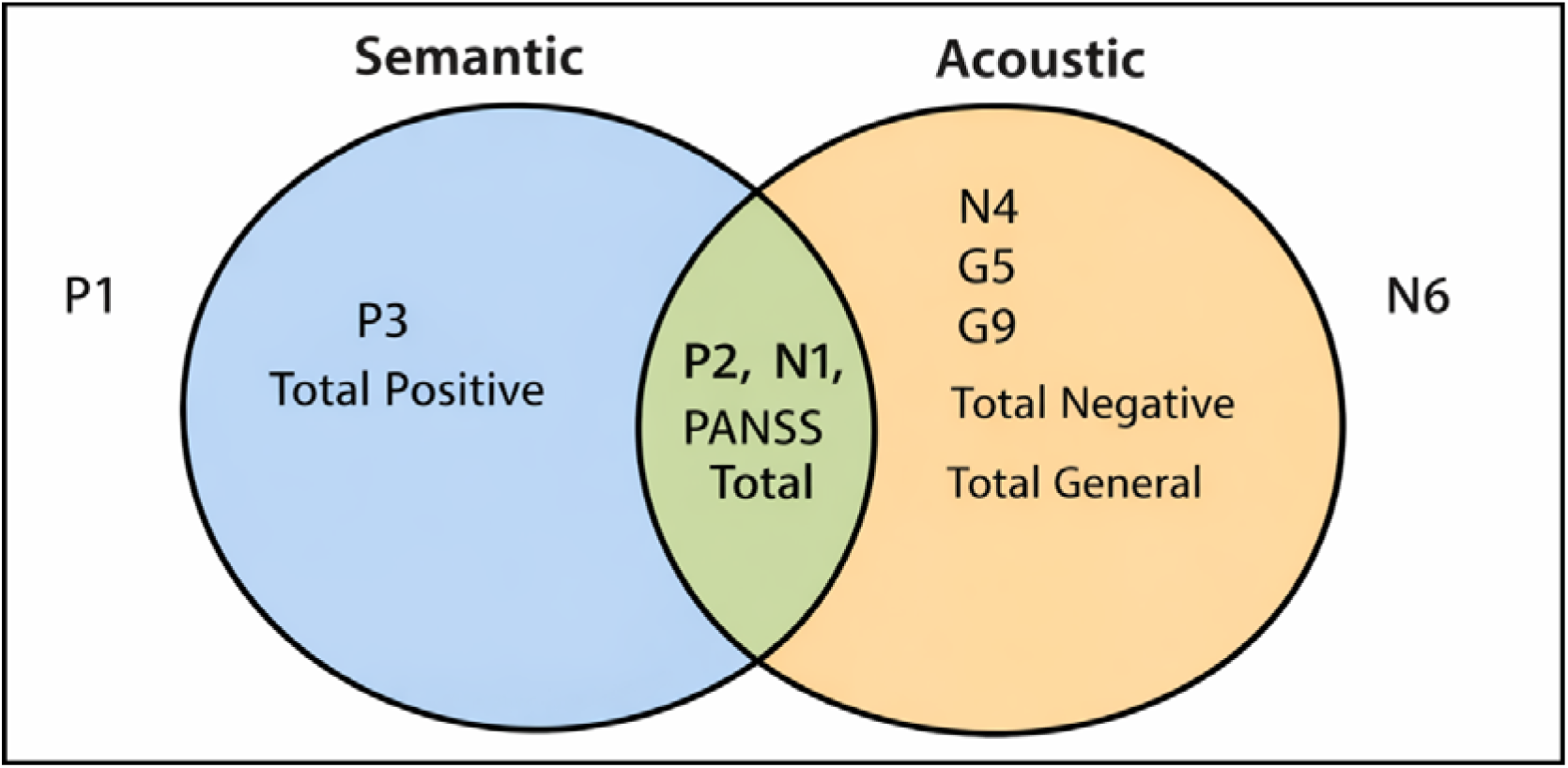
Venn diagram of shared and specific PANSS associations for semantic and acoustic indices.

### 3.4 Complementary and differential contribution of each index

Some PANSS scores were predicted by only one of the indices, whereas three items (P2, N1, PANSS Total) were significantly predicted by both of them. This suggests that the semantic and acoustic indices share some degree of predictive power for those dimensions. The relationship between both indices is illustrated in **Figure 2**, which confirms a significant but moderate correlation (Pearson’s). This indicates that although the indices partially overlap in the information they explain, each also provides differential contributions.

**Figure 2:**
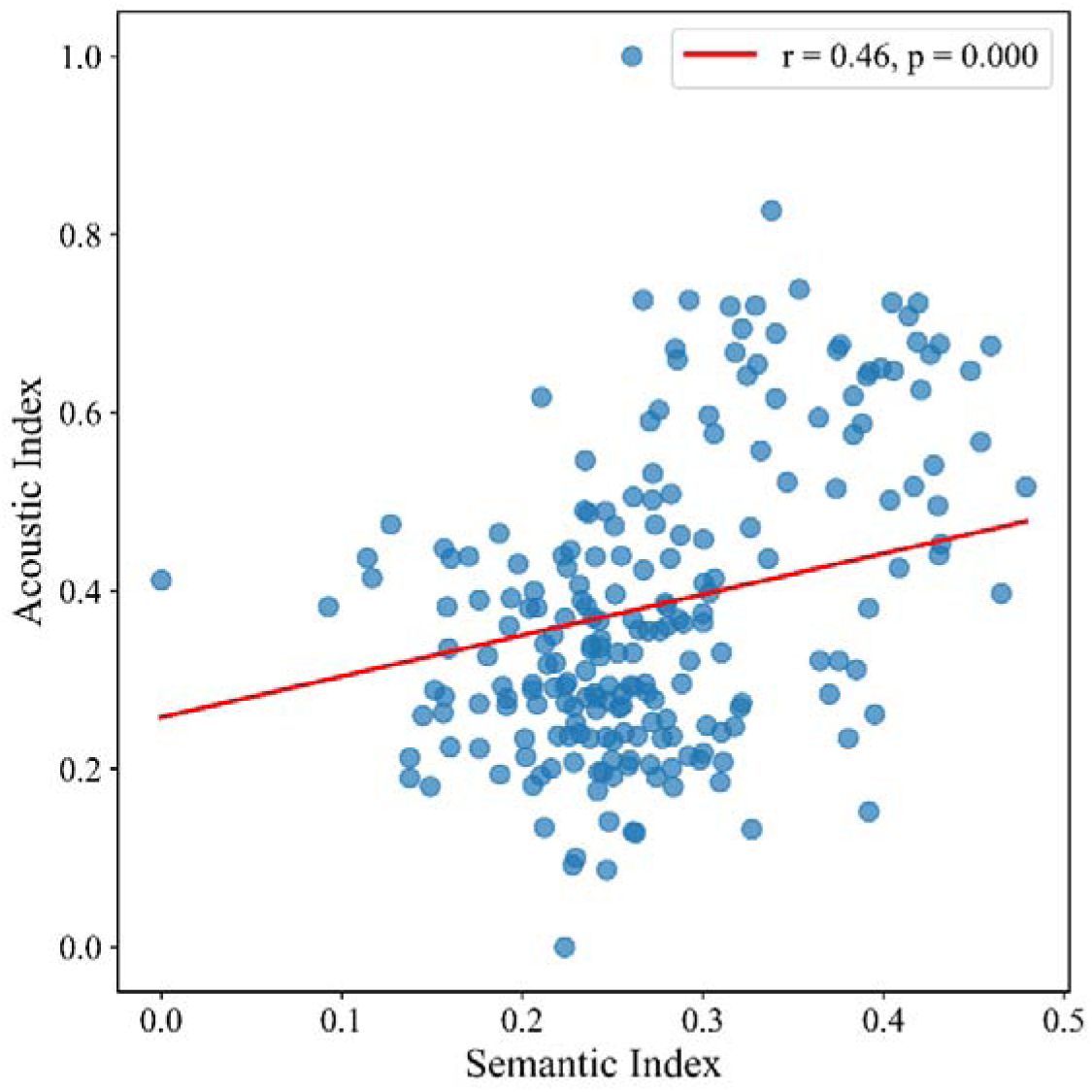
Correlation between semantic and acoustic indices.

The commonality analysis for predicting P2, N1 and PANSS Total using both indices is presented in **Table 6**. The results revealed distinct patterns of shared and differential contribution between the semantic and acoustic indices across symptoms. For P2, the semantic index accounted for the largest proportion of unique explained variance, with a small shared component between both indices. In contrast, N1 showed a stronger acoustic contribution, while retaining substantial shared variance and independent semantic contribution. Similarly, for PANSS Total, the acoustic index accounted for most of the unique explained variance, with a proportion of variance shared between both indices. Overall, these findings confirm the patterns observed across the remaining PANSS scores, in which the semantic index exhibited greater predictive power for positive symptoms, whereas the acoustic index showed stronger predictive power for the negative symptoms.

**Table 6:**
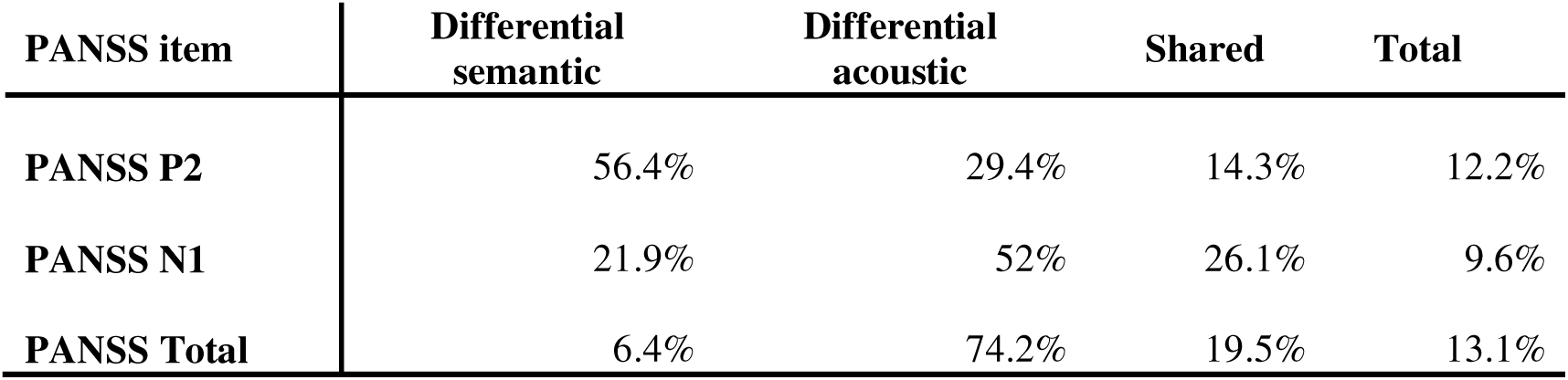
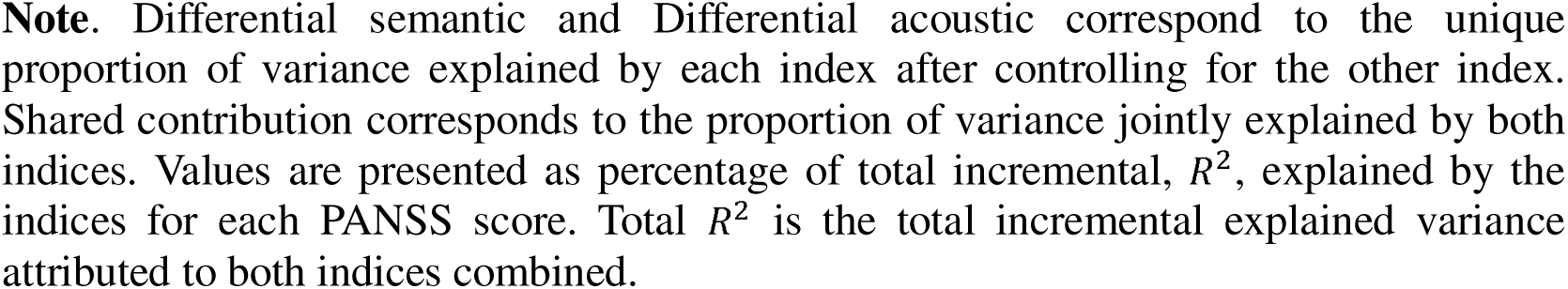
Shared and differential contribution of semantic and acoustic indices to PANSS scores.

| PANSS item | Differential semantic | Differential acoustic | Shared | Total |
| --- | --- | --- | --- | --- |
| PANSS P2 | 56.4% | 29.4% | 14.3% | 12.2% |
| PANSS N1 | 21.9% | 52% | 26.1% | 9.6% |
| PANSS Total | 6.4% | 74.2% | 19.5% | 13.1% |

**Note.** Differential semantic and Differential acoustic correspond to the unique proportion of variance explained by each index after controlling for the other index. Shared contribution corresponds to the proportion of variance jointly explained by both indices. Values are presented as percentage of total incremental, $R^2$ , explained by the indices for each PANSS score. Total $R^2$ is the total incremental explained variance attributed to both indices combined.

## 4. Discussion

Our findings extend previous work on a composite index by demonstrating that semantic and acoustic information can be mapped into parallel low-dimensional markers that capture distinct aspects of psychopathology. Building upon Palominos et al. (2025), we have constructed a semantic index based on text features extracted using embeddings from contextual and non-contextual language models, and an acoustic index based on 88 acoustic features extracted through eGeMAPS. Both indices proved to have predictive but also differential power. The semantic index was more strongly associated with positive symptoms, whereas the acoustic index mapped onto negative and general symptoms. At the same time, both indices contributed to the prediction of PANSS Total as well as the P2 (Conceptual Disorganization) and N1 (Blunted Affect) items, indicating that some symptom dimensions are represented across these two modalities. Together, these findings support the idea that complex speech features can be summarized into clinically meaningful latent dimensions.

More broadly, the results also support the idea that symptoms cannot be adequately characterized by a single speech-based index. Instead, different aspects of psychopathology seem to be reflected in partially overlapping but dissociable language domains. This observation is consistent with our initial motivation of developing a family of indices rather than a single omnibus metric. Such a family could track psychosis symptoms related to relapse or likelihood to remission by targeting specific dimensions relevant to this aim (Siafis et al., 2024). If speech disturbances emerge from distinct underlying mechanisms, then different representational spaces may be required to capture different symptom expressions. Thus, semantic and acoustic indices should be viewed as complementary markers of latent organization of language in psychopathology rather than competing alternatives.

Positive symptoms captured by semantic features primarily included several measures related to formal thought disorder and conceptual disorganization. This finding is consistent with previous literature showing that semantic embedding measures are sensitive to alterations in semantic organization and lexical selection in psychosis (Nour et al., 2023; Palominos et al., 2024; He et al., 2024). In contrast, acoustic features were more strongly associated with negative and general symptoms, including blunted affect (N1), passive apathetic social withdrawal (N4), mannerisms and posturing (G5), and unusual thought content (G9). These symptoms may manifest through changes in prosody, vocal expression, speech rate, pausing patterns, and other suprasegmental properties that are directly reflected in acoustic measurements.

Nevertheless, there was an association between the acoustic index and P2 (Conceptual Disorganization), a symptom traditionally linked to disturbances of thought and language organization. This result suggests that acoustic features may capture aspects of meaning indirectly through speech production mechanisms. Although semantic and acoustic information are often treated as separate domains, growing evidence indicates that they are partially intertwined. For example, Nygaard et al. (2009) demonstrated that speakers systematically use distinct prosodic patterns when producing words and that semantically related words tend to share acoustic characteristics. More recently, research on iconic prosody has shown that prosodic structure can contribute directly to meaning representation (Akita & Kawahara, 2025), while its close relationship with gesture suggests that speech acoustics may participate in broader multimodal systems of communication (Perlman, 2026). Consequently, acoustic measures may carry information that is not purely phonetic but also reflects aspects of semantic and communicative organization.

Our findings are also consistent with a recent study combining semantic and acoustic features. In a Dutch sample of 356 patients with SSD, Ciampelli et al. (2026) demonstrated that both acoustic and semantic measures were able to predict PANSS items. With combinations of different speech and language features, score on individual PANSS items can be predicted with < 1 point error. Importantly, they similarly observed that semantic features could best predict positive items, while acoustic features mapped closer to negative items. On the other hand, Ben Moshe et al. (2024) showed that acoustic prosodic features from a single spoken response can effectively detect schizophrenia symptoms and outperform textual features, with text providing only a slight additional improvement when combined. These studies converge on the conclusion that semantic and acoustic speech representations provide partially overlapping but distinct clinical information, with semantic features weighted towards positive symptoms and acoustic features towards negative and general symptoms.

Task-related effects also support this interpretation, since for the semantic index, task effects were observed primarily for positive symptoms, whereas associations with negative symptoms were comparatively stable across tasks, without observing task effects. This pattern seems to be plausible given that positive symptoms become more apparent in tasks with greater demand on discourse organization and semantic navigation. By contrast, negative symptoms may be reflected in reduced expressive behavior that remains relatively stable across different elicitation contexts. Consistent with this, for the acoustic index, task effects were more evident in free-speech, which imposes less structural constraints and allows greater expression of individual differences in speech production. Free speech may therefore provide a particularly sensitive condition for acoustic differences across symptoms.

The association between N1 (Blunted Affect) and the semantic index is somewhat less intuitive and expected, as blunted affect can be conceptualized as a reduction in emotional expression rather than a disturbance of semantic organization. However, diminished emotional engagement may also influence the content and structure of speech. Reduced affective expressiveness can lead to less elaborated discourse and lower variability in lexical choices, all of which may alter the semantic patterns captured by embedding-based measures. In this sense, the semantic index may not be detecting emotional expression directly, but rather downstream effects of affective flattening on language production. This interpretation is consistent with the broader view that semantic measures capture higher-order organizational properties of discourse, which can be influenced not only by positive symptoms but also by certain negative symptom dimensions.

In contrast, some symptom dimensions showed little or no relationship with either index. For example, P1 (Delusions) was not robustly associated with the semantic or acoustic indices. Delusions are fundamentally referential and belief-based phenomena, involving the interpretation of reality rather than necessarily altering the formal organization of speech. Consequently, their manifestation may be less detectable through measures of semantic structure or vocal expression alone. Capturing such symptoms may require a different type of analyses focused on referential content, narrative structure, pragmatic inference, or discourse-level representations.

### Limitations

Some limitations should be considered when interpreting the present findings. First, although the semantic and acoustic indices showed robust associations with symptom severity across datasets, the available samples were not fully balanced across languages, tasks, and participant characteristics. Composite indices are inherently data-driven, and their precise construction may be influenced by the relative representation of different subgroups. However, the fact that significant relations emerged despite considerable heterogeneity across datasets suggests that the indices capture signals of symptoms that are robust to variations in language, task, and sample composition. Future work using larger and more balanced datasets will be important to refine the indices, assess their stability across populations, and determine the extent to which the predictive performance generalizes.

Second, the methodology used to construct the indices is not unique. Alternative approaches for feature selection, weighting, dimensionality reduction, or index aggregation could produce somewhat different latent representations. The present study should therefore be viewed as one possible implementation within a broader framework of composite speech-based markers. Future studies should compare alternative index-construction strategies and evaluate their relative predictive performance and interpretability.

A further limitation is that our analyses focused on symptom dimensions selected because of their relevance to relapse prediction and longitudinal clinical monitoring. Consequently, we did not investigate the full range of PANSS items or other psychopathological dimensions. Additional work will be necessary to determine whether similar semantic and acoustic indices are associated with symptoms that were not examined here, and whether further symptom-specific indices can be developed.

Finally, the cross-linguistic nature of the study represents both a strength and a challenge. By combining data from five languages, we were able to evaluate whether we can generalize results across different linguistic contexts. However, differences in data collection procedures, sample composition, task administration, and clinical assessment introduce additional sources of variability. In contrast to Palominos et al. (2025), which followed the same cohort longitudinally, the present work relied primarily on cross-sectional data collected from different populations. Future studies with larger longitudinal datasets will be important to determine the temporal stability of the indices and their utility for tracking symptom trajectories within individuals.

## CRediT authorship contribution statement

**Maryia Kirdun**: Writing – review & editing, Writing – original draft, Visualization, Methodology, Investigation, Formal analysis. **Rui He**: Writing – review & editing, Methodology, Investigation. **Cemal Demirlek**: Data Curation. **José Tomás García-Molina**: Resources, Data Curation. **Roya Hüppi**: Data Curation. **Burcu Verim**: Data Curation. **Berna Yalincetin**: Data Curation. **Victor Ortiz García de la Foz**: Writing – review & editing, Resources, Data Curation. **Rosa Ayesa Arriola**: Writing – review & editing, Resources. **Emre Bora**: Writing – review & editing, Resources. **Alicia I. Figueroa-Barra**: Writing – review & editing, Resources. **Filip Spaniel**: Writing – review & editing, Resources. **Lena Palaniyappan**: Writing – review & editing, Resources, Funding acquisition. **Iris E. Sommer**: Writing – review & editing, Funding acquisition. **Philipp Homan**: Writing – review & editing, Funding acquisition. **Wolfram Hinzen**: Writing – review & editing, Writing – original draft, Supervision, Funding acquisition, Conceptualization. **Claudio Palominos**: Writing – review & editing, Writing – original draft, Project administration, Methodology, Investigation, Formal analysis, Conceptualization.

## Data Availability

All data produced in the present study are available upon reasonable request to the authors.

## Acknowledgments

This work is part of the TRUSTING project (TRUSTworthy speech-based AI monitorING system for the prediction of relapse in individuals with schizophrenia), funded by the European Union’s Horizon Europe research and innovation programme under grant agreement No 101080251. Data collection was also supported by the following local funding sources: Ministry of Health of the Czech Republic, grant nr. NU22-04-00143; National Agency for Research and Development (ANID), Chile (Fondecyt Regular Grant No. 1241618, to AFB); Swiss National Science Foundation (Grant No. 501100001711-191938, to ND), the Brain & Behaviour Research Foundation (Grant No. 28997, to WS), and the OPO Foundation (Grant No. 2020-0075, to WS and PH); Scientific and Technological Research Council of Turkey (TUBITAK 2247 Project No: 120C141). In addition, RAA was financed by a Miguel Servet contract from the Carlos III Health Institute (Grant No. CP18/00003) and a Consolidator Grant from the Ministerio de Ciencia e Innovación (Grant No. CNS2022-136110). LP acknowledges support from the Monique H. Bourgeois Chair and a salary award from the Fonds de recherche du Québec-Santé (FRQS: 366934) and a FRQS Research Centre Grant to the Douglas Research Centre (doi:10.69777/5230). The wide collaboration behind this work was made possible largely through the support of the DISCOURSE in Psychosis Consortium (https://discourseinpsychosis.org/), whose assistance was also instrumental in developing the speech assessment protocol for data collection at the separate sites.

## Notes

### Competing Interest Statement

Lena Palaniyappan reports personal fees for serving as chief editor from the Canadian Medical Association Journals, speaker/consultant fee from Janssen Canada and Otsuka Canada, SPMM Course Limited, UK, Canadian Psychiatric Association; book royalties from Oxford University Press; investigator-initiated educational grants from Janssen Canada, Sunovion and Otsuka Canada outside the submitted work. Iris E. Sommer reports charity grant from Janssen, speaker fee from Otzuka and Ludbeck. Philipp Homan has received grants and honoraria from Novartis, Lundbeck, Takeda, Mepha, Janssen, Boehringer Ingelheim, Neurolite and OM Pharma outside of this work. All other authors reported no conflict of interests.

### Author Declarations

Cezch: The Ethics Committee of the National Institute of Mental Health, Czech Republic, gave ethical approval for this work. Spanish: The Research Ethics Committee for Medicinal Products and Medical Devices of Cantabria, Cantabrian Health Service, gave ethical approval for this work at the Valdecilla Research Institute and Marques de Valdecilla University Hospital. Chilean-Spanish: The Scientific Ethics Committee of the Barros Luco Healthcare Complex, within the Metropolitan South Health Service, gave ethical approval for this work at Barros Luco Trudeau Clinical Hospital. Swiss-German: The Cantonal Ethics Committee Zurich gave ethical approval for this work at the Department of Adult Psychiatry and Psychotherapy, Psychiatric Hospital of the University of Zurich. Turkish: The Ethics Committee of Dokuz Eylul University gave ethical approval for this work at the Psychotic Disorders Outpatient Unit, Department of Psychiatry, Dokuz Eylul University.

## References

Agurto, C., Pietrowicz, M., Norel, R., Eyigoz, E. K., Stanislawski, E., Cecchi, G., & Corcoran, C. (2020). Analyzing acoustic and prosodic fluctuations in free speech to predict psychosis onset in high-risk youths. Annual International Conference of the IEEE Engineering in Medicine and Biology Society. IEEE Engineering in Medicine and Biology Society. Annual International Conference, 2020, 5575–5579. 10.1109/EMBC44109.2020.9176841.

Akita, K., & Kawahara, S. (2025). Iconic prosody enhances the depictive power of ideophones. Language and Cognition, 17, e77.

Alonso-Sánchez, M. F., Limongi, R., Gati, J., & Palaniyappan, L. (2023). Language network self-inhibition and semantic similarity in first-episode schizophrenia: A computational-linguistic and effective connectivity approach. Schizophrenia research, 259, 97–103.

Arslan, B., Kizilay, E., Verim, B., Demirlek, C., Dokuyan, Y., Turan, Y. E., Kucukakdag, A., Demir, M., Cesim, E., & Bora, E. (2024). Automated linguistic analysis in speech samples of Turkish-speaking patients with schizophrenia-spectrum disorders. Schizophrenia research, 267, 65–71. 10.1016/j.schres.2024.03.014.

Bar, K., Zilberstein, V., Ziv, I., Baram, H., Dershowitz, N., Itzikowitz, S., & Harel, E. V. (2019). Semantic characteristics of schizophrenic speech. In Proceedings of the Sixth Workshop on Computational Linguistics and Clinical Psychology (pp. 84–93). Association for Computational Linguistics.

Bedi, G., Carrillo, F., Cecchi, G. A., Slezak, D. F., Sigman, M., Mota, N. B.,…& Corcoran, C. M. (2015). Automated analysis of free speech predicts psychosis onset in high-risk youths. npj Schizophrenia, 1(1), 1–7.

Ben Moshe, T., Ziv, I., Dershowitz, N., & Bar, K. (2024). The contribution of prosody to machine classification of schizophrenia. Schizophrenia, 10(1), 53.

Berardi, M., Brosch, K., Pfarr, J. K., Schneider, K., Sültmann, A., Thomas-Odenthal, F., Wroblewski, A., Usemann, P., Philipsen, A., Dannlowski, U., Nenadić, I., Kircher, T., Krug, A., Stein, F., & Dietrich, M. (2023). Relative importance of speech and voice features in the classification of schizophrenia and depression. Translational psychiatry, 13(1), 298. 10.1038/s41398-023-02594-0

Buchanan, R. W., Davidson, M.,…Leucht, S. (2024). Relapse in clinically stable adult patients with schizophrenia or schizoaffective disorder: evidence-based criteria derived by equipercentile linking and diagnostic test accuracy meta-analysis. The lancet. Psychiatry, 11(1), 36–46. 10.1016/S2215-0366(23)00364-4

Chakrabartty, S. N. (2017). Composite index: methods and properties. Journal of Applied Quantitative Methods, 12(2), 25–33.

Chao, Y. S., & Wu, C. J. (2017). Principal component-based weighted indices and a framework to evaluate indices: Results from the Medical Expenditure Panel Survey 1996 to 2011. PLoS One, 12(9), e0183997.

Ciampelli, S., de Boer, J. N., Koops, S., Troelstra, E., Jebens, A., Marsman, J. B. C.,…& Sommer, I. E. (2026). Automated speech-based modeling of item-level symptom severity in schizophrenia. JAMA Network Open, 9(6), e2620239.

Çokal, D., Aloraini, A., Palominos, C. F., Demirlek, C., Verim, B., Yalınçetin, B.,…& Hinzen, W. (2025). Three dimensions of speech coherence in people with early psychosis and their family members. Schizophrenia.

da Silva, A. C. B., dos Reis, D. R., Libório, M. P., Mannan, H., & Nobre, C. N. (2026). A Composite Indicator of Anxiety and Depression Signs in Children and Adolescents of Low-and Middle-Income Countries: a Subjective-Objective Multidimensional Approach. Child Indicators Research, 1–40.

De Boer, J. N., Voppel, A. E., Brederoo, S. G., Schnack, H. G., Truong, K. P., Wijnen, F. N. K., & Sommer, I. E. C. (2023). Acoustic speech markers for schizophrenia-spectrum disorders: a diagnostic and symptom-recognition tool. Psychological medicine, 53(4), 1302–1312.

Eyben, F., Wöllmer, M., & Schuller, B. (2010). openSMILE: The Munich versatile and fast open-source audio feature extractor. In Proceedings of the 18th ACM International Conference on Multimedia (pp. 1459–1462). Association for Computing Machinery. 10.1145/1873951.1874246

Eyben, F., Weninger, F., Gross, F., & Schuller, B. (2013). Recent developments in openSMILE, the Munich open-source multimedia feature extractor. In Proceedings of the 21st ACM International Conference on Multimedia (pp. 835–838). Association for Computing Machinery. 10.1145/2502081.2502224

Eyben, F., Scherer, K. R., Schuller, B. W., Sundberg, J., André, E., Busso, C., Devillers, L., Epps, J., Laukka, P., Narayanan, S., & Thuong, K. (2016). The Geneva Minimalistic Acoustic Parameter Set (GeMAPS) for voice research and affective computing. IEEE Transactions on Affective Computing, 7(2), 190–202. 10.1109/TAFFC.2015.2457417.

Figueroa-Barra, A., Del Aguila, D., Cerda, M., Gaspar, P. A., Terissi, L. D., Durán, M., & Valderrama, C. (2022). Automatic language analysis identifies and predicts schizophrenia in first-episode of psychosis. Schizophrenia, 8(1), 53.

García-Gutiérrez F., Marquié M., Muñoz N., Alegret M., Cano A., de Rojas I., García-González P., Olivé C., Puerta R., Orellana A., Montrreal L., Pytel V., Ricciardi M., Zaldua C., Gabirondo P., Hinzen W., Lleonart N., García-Sánchez A., Tárraga L., Ruiz A., Boada M. and Valero S. (2023). Harnessing acoustic speech parameters to decipher amyloid status in individuals with mild cognitive impairment. Front. Neurosci. 17:1221401. doi: 10.3389/fnins.2023.1221401

Grot, S., Giguère, C.-É., Smine, S., Mongeau, V., Nguyen, D., Preda, A., Potvin, S., Erp, T. G. M. van, FBIRN, & Orban, P. (2021). Converting scores between the PANSS and SAPS/SANS beyond the positive/negative dichotomy. Psychiatry Research, 305, 114199. 10.1016/j.psychres.2021.114199

Haider, F., de la Fuente, S., & Luz, S. (2020). An assessment of paralinguistic acoustic features for detection of Alzheimer’s dementia in spontaneous speech. IEEE Journal of Selected Topics in Signal Processing, 14(2), 272–281. 10.1109/JSTSP.2019.2955022

He, R., Chapin, K., Al-Tamimi, J., Bel, N., Marquié, M., Rosende-Roca, M.,…& Hinzen, W. (2023). Automated classification of cognitive decline and probable Alzheimer’s dementia across multiple speech and language domains. American Journal of Speech-Language Pathology, 32(5), 2075–2086.

He, R., Palominos, C., Zhang, H., Alonso-Sánchez, M. F., Palaniyappan, L., & Hinzen, W. (2024). Navigating the semantic space: Unraveling the structure of meaning in psychosis using different computational language models. Psychiatry Research, 333, 115752.

He, R., Kirdun, M., Palominos, C., Orejudo, L. N., Barthelemy, S., Bhola, S.,…& Hinzen, W. (2026). Predicting PANSS symptoms in schizophrenia spectrum disorders using speech only: an international, multi-centre, retrospective, computational study across multiple languages. medRxiv, 2026-02.

Horn, J.L. (1965). A rationale and test for the number of factors in factor analysis. Psychometrica 30: 179–185.

Hüppi, R. M., Surbeck, W., Pauli, Y. L., Dannecker, N., Fabian, D., Edkins, V.,…& Homan, P. (2026). Dissociable computational markers of semantic search and verbal retrieval drive across the psychosis spectrum. medRxiv, 2026-05.

Longman, R. S., Cota, A. A., Holden, R. R., & Fekken, G. C. (1989). A Regression Equation for the Parallel Analysis Criterion in Principal Components Analysis: Mean and 95th Percentile Eigenvalues. Multivariate Behavioral Research, 24(1), 59–69. 10.1207/s15327906mbr2401_4

Melshin, G., DiMaggio, A., Zeramdini, N., MacKinley, M., Palaniyappan, L., & Voppel, A. (2025). Taking a look at your speech: identifying diagnostic status and negative symptoms of psychosis using convolutional neural networks. NPP—Digital Psychiatry and Neuroscience, 3(1), 19.

Meta AI. (2024). The LLaMA 3 herd of models. https://ai.meta.com/research/publications/the-llama-3-herd-of-models/

Nardo, M., Saisana, M., Saltelli, A., & Tarantola, S. (2005). Tools for composite indicators building. *European Comission*, Ispra, 15(1), 19–20.

Nour, M. M., McNamee, D. C., Liu, Y., & Dolan, R. J. (2023). Trajectories through semantic spaces in schizophrenia and the relationship to ripple bursts. Proceedings of the National Academy of Sciences, 120(42), e2305290120.

Nygaard, L. C., Herold, D. S., & Namy, L. L. (2009). The semantics of prosody: Acoustic and perceptual evidence of prosodic correlates to word meaning. Cognitive science, 33(1), 127–146.

Palominos, C., He, R., Fröhlich, K., Mülfarth, R. R., Seuffert, S., Sommer, I. E.,…& Hinzen, W. (2024). Approximating the semantic space: word embedding techniques in psychiatric speech analysis. Schizophrenia, 10(1), 114.

Palominos, C., Kirdun, M., Nikzad, A. H., Spilka, M. J., Homan, P., Sommer, I. E.,…& Hinzen, W. (2025). A single composite index of semantic behavior tracks symptoms of psychosis over time. Schizophrenia Research, 279, 116–127.

Perlman, M. (2026). Iconic prosody and its connection to iconic gesture.

Pintos, A. S., Hui, C. L. M., De Deyne, S., Cheung, C., Ko, W. T., Nam, S. Y.,…& Chen, E. Y. H. (2022). A longitudinal study of semantic networks in schizophrenia and other psychotic disorders using the word association task. Schizophrenia Bulletin Open, 3(1), sgac054.

Pokorny, F. B., Schmitt, M., Egger, M., Bartl-Pokorny, K. D., Zhang, D., Schuller, B. W., & Marschik, P. B. (2022). Automatic vocalisation-based detection of fragile X syndrome and Rett syndrome. Scientific reports, 12(1), 13345. 10.1038/s41598-022-17203-1

R Core Team (2025). *R: A Language and Environment for Statistical Computing*. R Foundation for Statistical Computing, Vienna, Austria. <https://www.R-project.org/>.

Rohanian, M., Hüppi, R., Nooralahzadeh, F., Dannecker, N., Pauli, Y., Surbeck, W.,…& Homan, P. (2026). Uncertainty modeling in multimodal speech analysis across the psychosis spectrum. npj Digital Medicine. doi: 10.1038/s41746-025-02309-3.

Siafis, S., Brandt, L., McCutcheon, R. A., Gutwinski, S., Schneider-Thoma, J., Bighelli, I.,…& Leucht, S. (2024). Relapse in clinically stable adult patients with schizophrenia or schizoaffective disorder: evidence-based criteria derived by equipercentile linking and diagnostic test accuracy meta-analysis. The Lancet Psychiatry, 11(1), 36–46.

Surbeck, W., Omlor, W., Dannecker, N., Samuel, R., Steiner, A., Fabian, D.,…& Homan, P. (2025). Altered white matter microstructure of language pathways and semantic cognition deficiencies in early psychosis. Schizophrenia, 11(1), 136. doi: 10.1038/s41537-025-00682-2.

Tang, S. X., Spilka, M. J., John, M., Birnbaum, M. L., Saito, E., Berretta, S. A.,…& Kane, J. M. (2025). Automated speech and language markers of longitudinal changes in psychosis symptoms. NPP—Digital Psychiatry and Neuroscience, 3(1), 13.

Teixeira, F. L., Costa, M. R. E., Abreu, J. P., Cabral, M., Soares, S. P., & Teixeira, J. P. (2023). A narrative review of speech and EEG features for Schizophrenia detection: Progress and challenges. Bioengineering, 10(4), 493.

Voppel, A. E., De Boer, J. N., Brederoo, S. G., Schnack, H. G., & Sommer, I. E. (2023). Semantic and acoustic markers in schizophrenia-spectrum disorders: A combinatory machine learning approach. Schizophrenia bulletin, 49(Supplement_2), S163–S171.

Worthington, M., Efstathiadis, G., Yadav, V., Galatzer-Levy, I., Kott, A., Pintilii, E.,…& Abbas, A. (2025). Measurement of schizophrenia symptoms through speech analysis from PANSS interview recordings. Frontiers in Psychiatry, 16, 1571647.

