## Supplemental Material for "A double index for assessing symptoms in psychosis based on acoustic and semantic information"

**Table S1**: Inclusion and exclusion criteria across sites.

| **Language** | **Stage of psychosis** | **Outpatients/inpatients** | **Inclusion Criteria** | **Exclusion Criteria** |
| --- | --- | --- | --- | --- |
| **Czech** | First episode (<2 years since onset) or multiple episodes with cumulative illness duration ≤1.5 years | Outpatients/inpatients | Age 18–60; diagnosis of schizophrenia (ICD-10 F20.X), acute polymorphic psychotic disorder with schizophrenia symptoms (F23.1), or acute schizophrenia-like psychotic disorder (F23.2) | Organic mental disorders, IQ < 80, severe neurological disease, metal implants in head/face, cardiac pacemaker |
| **Spanish** | Chronic schizophrenia patients | Outpatients | DSM-IV diagnosis of brief psychotic disorder, schizophreniform disorder, schizophrenia, or schizoaffective disorder | Meeting DSM-IV criteria for drug dependence or mental retardation; history of neurological disease or head injury |
| **Chilean-Spanish** | First episode | Outpatients | DSM-IV diagnosis of brief psychotic disorder, schizophreniform disorder, schizophrenia, or schizoaffective disorder | Medical or neurological disorders; current alcohol/substance abuse |
| **Swiss-German** | Early psychosis | Outpatients/inpatients | ICD-10 psychotic disorders; age 14–40; within first 8 years of psychosis onset; right-handed; admitted to Psychiatric University Clinic Zurich | Substance intoxication/withdrawal (including cannabis ≥1 month), ophthalmological or neurological conditions, head trauma, history of loss of consciousness, pregnancy |
| **Turkish** | 35 first-episode psychosis, 56 chronic schizophrenia, and 17 chronic schizoaffective patients. | Outpatient | Native Turkish speakers; DSM-IV schizophrenia spectrum disorder diagnosed using SCID-I | Medical or neurological disorders; current alcohol/substance abuse |

**Table S2**: List of all acoustic eGeMAPS features.

| **Fundamental Frequency (F0)** | |
| --- | --- |
| 1 | F0semitoneFrom27.5Hz_sma3nz_amean |
| 2 | F0semitoneFrom27.5Hz_sma3nz_stddevNorm |
| 3 | F0semitoneFrom27.5Hz_sma3nz_percentile20.0 |
| 4 | F0semitoneFrom27.5Hz_sma3nz_percentile50.0 |
| 5 | F0semitoneFrom27.5Hz_sma3nz_percentile80.0 |
| 6 | F0semitoneFrom27.5Hz_sma3nz_pctlrange0-2 |
| 7 | F0semitoneFrom27.5Hz_sma3nz_meanRisingSlope |
| 8 | F0semitoneFrom27.5Hz_sma3nz_stddevRisingSlope |
| 9 | F0semitoneFrom27.5Hz_sma3nz_meanFallingSlope |
| 10 | F0semitoneFrom27.5Hz_sma3nz_stddevFallingSlope |
| **Loudness** | |
| 11 | loudness_sma3_amean |
| 12 | loudness_sma3_stddevNorm |
| 13 | loudness_sma3_percentile20.0 |
| 14 | loudness_sma3_percentile50.0 |
| 15 | loudness_sma3_percentile80.0 |
| 16 | loudness_sma3_pctlrange0-2 |
| 17 | loudness_sma3_meanRisingSlope |
| 18 | loudness_sma3_stddevRisingSlope |
| 19 | loudness_sma3_meanFallingSlope |
| 20 | loudness_sma3_stddevFallingSlope |
| 21 | loudnessPeaksPerSec |
| 22 | equivalentSoundLevel_dBp |
| **Spectral Features** | |
| 23 | alphaRatioV_sma3nz_amean |
| 24 | alphaRatioV_sma3nz_stddevNorm |
| 25 | alphaRatioUV_sma3nz_amean |
| 26 | hammarbergIndexV_sma3nz_amean |
| 27 | hammarbergIndexV_sma3nz_stddevNorm |
| 28 | hammarbergIndexUV_sma3nz_amean |
| 29 | slopeV0-500_sma3nz_amean |
| 30 | slopeV0-500_sma3nz_stddevNorm |
| 31 | slopeV500-1500_sma3nz_amean |
| 32 | slopeV500-1500_sma3nz_stddevNorm |
| 33 | slopeUV0-500_sma3nz_amean |
| 34 | slopeUV500-1500_sma3nz_amean |
| 35 | spectralFluxV_sma3nz_amean |
| 36 | spectralFluxV_sma3nz_stddevNorm |
| 37 | spectralFluxUV_sma3nz_amean |
| 38 | spectralFlux_sma3_amean |
| 39 | spectralFlux_sma3_stddevNorm |
| **Formants (F1–F3)** | |
| 40 | F1frequency_sma3nz_amean |
| 41 | F1frequency_sma3nz_stddevNorm |
| 42 | F1bandwidth_sma3nz_amean |
| 43 | F1bandwidth_sma3nz_stddevNorm |
| 44 | F1amplitudeLogRelF0_sma3nz_amean |
| 45 | F1amplitudeLogRelF0_sma3nz_stddevNorm |
| 46 | F2frequency_sma3nz_amean |
| 47 | F2frequency_sma3nz_stddevNorm |
| 48 | F2bandwidth_sma3nz_amean |
| 49 | F2bandwidth_sma3nz_stddevNorm |
| 50 | F2amplitudeLogRelF0_sma3nz_amean |
| 51 | F2amplitudeLogRelF0_sma3nz_stddevNorm |
| 52 | F3frequency_sma3nz_amean |
| 53 | F3frequency_sma3nz_stddevNorm |
| 54 | F3bandwidth_sma3nz_amean |
| 55 | F3bandwidth_sma3nz_stddevNorm |
| 56 | F3amplitudeLogRelF0_sma3nz_amean |
| 57 | F3amplitudeLogRelF0_sma3nz_stddevNorm |
| **MFCC (Mel-Frequency Cepstral Coefficients)** | |
| 58 | mfcc1_sma3_amean |
| 59 | mfcc1_sma3_stddevNorm |
| 60 | mfcc1V_sma3nz_amean |
| 61 | mfcc1V_sma3nz_stddevNorm |
| 62 | mfcc2_sma3_amean |
| 63 | mfcc2_sma3_stddevNorm |
| 64 | mfcc2V_sma3nz_amean |
| 65 | mfcc2V_sma3nz_stddevNorm |
| 66 | mfcc3_sma3_amean |
| 67 | mfcc3_sma3_stddevNorm |
| 68 | mfcc3V_sma3nz_amean |
| 69 | mfcc3V_sma3nz_stddevNorm |
| 70 | mfcc4_sma3_amean |
| 71 | mfcc4_sma3_stddevNorm |
| 72 | mfcc4V_sma3nz_amean |
| 73 | mfcc4V_sma3nz_stddevNorm |
| **Voice Quality** | |
| 74 | jitterLocal_sma3nz_amean |
| 75 | jitterLocal_sma3nz_stddevNorm |
| 76 | shimmerLocaldB_sma3nz_amean |
| 77 | shimmerLocaldB_sma3nz_stddevNorm |
| 78 | HNRdBACF_sma3nz_amean |
| 79 | HNRdBACF_sma3nz_stddevNorm |
| 80 | logRelF0-H1-H2_sma3nz_amean |
| 81 | logRelF0-H1-H2_sma3nz_stddevNorm |
| 82 | logRelF0-H1-A3_sma3nz_amean |
| 83 | logRelF0-H1-A3_sma3nz_stddevNorm |
| **Voicing & Temporal** | |
| 84 | VoicedSegmentsPerSec |
| 85 | MeanVoicedSegmentLengthSec |
| 86 | StddevVoicedSegmentLengthSec |
| 87 | MeanUnvoicedSegmentLength |
| 88 | StddevUnvoicedSegmentLength |

**Table S3**: Linear regression results predicting PANSS scores using semantic an acoustic indices.

| **Semantic Index** | |  |  |  |  |  |  |  |
| --- | --- | --- | --- | --- | --- | --- | --- | --- |
| **Model** | **Term** | **β** | **SE** | **t** | **p** | **Sig.** | **R²** | **adj. R²** |
| **P1** | **log(geo_mean_index)** | 0.357 | 0.271 | 1.319 | 0.189 | — | 0.026 | 0.012 |
|  | Age | -0.007 | 0.007 | -0.976 | 0.33 | — |  |  |
|  | Sex (male) | 0.172 | 0.155 | 1.115 | 0.266 | — |  |  |
| **P2** | **log(geo_mean_index)** | 0.932 | 0.247 | 3.773 | <0.001 | ******* | 0.109 | 0.097 |
|  | Age | -0.01 | 0.007 | -1.557 | 0.121 | — |  |  |
|  | Sex (male) | 0.218 | 0.141 | 1.549 | 0.123 | — |  |  |
| **P3** | **log(geo_mean_index)** | 0.67 | 0.272 | 2.462 | 0.015 | ***** | 0.09 | 0.077 |
|  | Age | -0.018 | 0.007 | -2.48 | 0.014 | ***** |  |  |
|  | Sex (male) | 0.212 | 0.155 | 1.364 | 0.174 | — |  |  |
| **N1** | **log(geo_mean_index)** | 1.072 | 0.371 | 2.886 | 0.005 | ****** | 0.18 | 0.158 |
|  | Age | -0.012 | 0.014 | -0.882 | 0.38 | — |  |  |
|  | Sex (male) | 0.557 | 0.267 | 2.084 | 0.04 | ***** |  |  |
| **N4** | **log(geo_mean_index)** | 0.194 | 0.359 | 0.542 | 0.589 | — | 0.163 | 0.139 |
|  | Age | -0.041 | 0.013 | -3.072 | 0.003 | ****** |  |  |
|  | Sex (male) | 0.261 | 0.258 | 1.011 | 0.314 | — |  |  |
| **N6** | **log(geo_mean_index)** | 0.016 | 0.342 | 0.047 | 0.963 | — | 0.191 | 0.169 |
|  | Age | -0.044 | 0.013 | -3.448 | <0.001 | ******* |  |  |
|  | Sex (male) | 0.375 | 0.246 | 1.524 | 0.13 | — |  |  |
| **G5** | **log(geo_mean_index)** | 0.012 | 0.209 | 0.057 | 0.954 | — | 0.09 | 0.065 |
|  | Age | -0.02 | 0.008 | -2.64 | 0.009 | ****** |  |  |
|  | Sex (male) | 0.001 | 0.15 | 0.007 | 0.994 | — |  |  |
| **G9** | **log(geo_mean_index)** | 0.284 | 0.299 | 0.948 | 0.345 | — | 0.152 | 0.128 |
|  | Age | -0.033 | 0.011 | -2.971 | 0.004 | ****** |  |  |
|  | Sex (male) | 0.064 | 0.215 | 0.296 | 0.768 | — |  |  |
| **Total Positive** | **log(geo_mean_index)** | 1.959 | 0.615 | 3.183 | 0.002 | ****** | 0.108 | 0.096 |
|  | Age | -0.035 | 0.016 | -2.151 | 0.033 | ***** |  |  |
|  | Sex (male) | 0.603 | 0.351 | 1.716 | 0.088 | — |  |  |
| **Total Negative** | **log(geo_mean_index)** | 1.282 | 0.927 | 1.383 | 0.169 | — | 0.209 | 0.188 |
|  | Age | -0.097 | 0.034 | -2.813 | 0.006 | ****** |  |  |
|  | Sex (male) | 1.192 | 0.667 | 1.788 | 0.077 | — |  |  |
| **Total General** | **log(geo_mean_index)** | 0.296 | 0.43 | 0.688 | 0.493 | — | 0.165 | 0.142 |
|  | Age | -0.053 | 0.016 | -3.353 | 0.001 | ****** |  |  |
|  | Sex (male) | 0.065 | 0.309 | 0.21 | 0.834 | — |  |  |
| **PANSS Total** | **log(geo_mean_index)** | 3.477 | 1.793 | 1.939 | 0.055 | . | 0.247 | 0.227 |
|  | Age | -0.211 | 0.066 | -3.176 | 0.002 | ****** |  |  |
|  | Sex (male) | 1.784 | 1.289 | 1.384 | 0.169 | — |  |  |

| **Acoustic Index** | |  |  |  |  |  |  |  |
| --- | --- | --- | --- | --- | --- | --- | --- | --- |
| **Model** | **Term** | **β** | **SE** | **t** | **p** | **Sig.** | **R²** | **adj. R²** |
| **P1** | **log(geo_mean_index)** | -0.318 | 0.256 | -1.243 | 0.215 | — | 0.025 | 0.011 |
|  | Sex (male) | 0.109 | 0.163 | 0.669 | 0.504 | — |  |  |
|  | Age | -0.007 | 0.007 | -1.024 | 0.307 | — |  |  |
| **P2** | **log(geo_mean_index)** | -0.912 | 0.233 | -3.923 | <0.001 | ******* | 0.114 | 0.102 |
|  | Sex (male) | 0.037 | 0.149 | 0.249 | 0.804 | — |  |  |
|  | Age | -0.01 | 0.007 | -1.59 | 0.113 | — |  |  |
| **P3** | **log(geo_mean_index)** | -0.034 | 0.26 | -0.131 | 0.896 | — | 0.065 | 0.052 |
|  | Sex (male) | 0.21 | 0.166 | 1.264 | 0.208 | — |  |  |
|  | Age | -0.023 | 0.007 | -3.164 | 0.002 | ****** |  |  |
| **N1** | **log(geo_mean_index)** | -1.015 | 0.447 | -2.272 | 0.025 | ***** | 0.158 | 0.134 |
|  | Sex (male) | 0.347 | 0.276 | 1.26 | 0.21 | — |  |  |
|  | Age | -0.013 | 0.015 | -0.906 | 0.367 | — |  |  |
| **N4** | **log(geo_mean_index)** | -1.191 | 0.41 | -2.903 | 0.004 | ****** | 0.221 | 0.199 |
|  | Sex (male) | 0.09 | 0.253 | 0.356 | 0.722 | — |  |  |
|  | Age | -0.023 | 0.013 | -1.679 | 0.096 | — |  |  |
| **N6** | **log(geo_mean_index)** | -0.848 | 0.397 | -2.135 | 0.035 | ***** | 0.223 | 0.202 |
|  | Sex (male) | 0.262 | 0.245 | 1.068 | 0.288 | — |  |  |
|  | Age | -0.028 | 0.013 | -2.185 | 0.031 | ***** |  |  |
| **G5** | **log(geo_mean_index)** | -0.721 | 0.238 | -3.027 | 0.003 | ****** | 0.16 | 0.137 |
|  | Sex (male) | -0.095 | 0.147 | -0.643 | 0.522 | — |  |  |
|  | Age | -0.007 | 0.008 | -0.956 | 0.341 | — |  |  |
| **G9** | **log(geo_mean_index)** | -1.36 | 0.332 | -4.098 | <0.001 | ******* | 0.259 | 0.238 |
|  | Sex (male) | -0.135 | 0.205 | -0.66 | 0.511 | — |  |  |
|  | Age | -0.013 | 0.011 | -1.218 | 0.226 | — |  |  |
| **Total Positive** | **log(geo_mean_index)** | -1.264 | 0.588 | -2.15 | 0.033 | ***** | 0.086 | 0.073 |
|  | Sex (male) | 0.357 | 0.376 | 0.949 | 0.344 | — |  |  |
|  | Age | -0.041 | 0.017 | -2.475 | 0.014 | ***** |  |  |
| **Total Negative** | **log(geo_mean_index)** | -3.054 | 1.07 | -2.854 | 0.005 | ****** | 0.251 | 0.231 |
|  | Sex (male) | 0.7 | 0.661 | 1.059 | 0.292 | — |  |  |
|  | Age | -0.064 | 0.035 | -1.833 | 0.07 | — |  |  |
| **Total General** | **log(geo_mean_index)** | -2.081 | 0.47 | -4.423 | <0.001 | ******* | 0.289 | 0.27 |
|  | Sex (male) | -0.23 | 0.29 | -0.791 | 0.431 | — |  |  |
|  | Age | -0.021 | 0.015 | -1.343 | 0.182 | — |  |  |
| **PANSS Total** | **log(geo_mean_index)** | -8.225 | 2.015 | -4.082 | <0.001 | ******* | 0.325 | 0.306 |
|  | Sex (male) | 0.458 | 1.244 | 0.368 | 0.714 | — |  |  |
|  | Age | -0.124 | 0.066 | -1.882 | 0.063 | — |  |  |

**Note**. Linear regression models were fitted with PANSS score as dependent variable and the log-transformed geometric mean index as the main predictor, controlling for age and sex. The sample size was N = 221 for models involving P-scale items and N = 113 for models involving N-scale, G-scale, and total PANSS scores, due to 108 observations with missing data being excluded from these analyses. *** p < .001, ** p < .01, * p < .05.

**Table S4**: Mixed-effects results predicting indices scores from task-level semantic and acoustic indices.

| **Semantic Index** |  |  |  |  |  |
| --- | --- | --- | --- | --- | --- |
| **Term** | **β** | **SE** | **z** | **p** | **Sig.** |
| **P1** | **0.027** | **0.021** | **1.241** | **0.215** | — |
| Age | -0.009 | 0.002 | -4.295 | <0.001 | ******* |
| Sex (male) | 0.005 | 0.049 | 0.092 | 0.926 | — |
| Task: fs | -0.207 | 0.043 | -4.819 | <0.001 | ******* |
| Task: pd | -0.192 | 0.037 | -5.178 | <0.001 | ******* |
| Task: story | -0.474 | 0.051 | -9.374 | <0.001 | ******* |
| **P2** | **0.078** | **0.022** | **3.493** | **<0.001** | ******* |
| Age | -0.008 | 0.002 | -3.885 | <0.001 | ******* |
| Sex (male) | -0.008 | 0.048 | -0.167 | 0.867 | — |
| Task: fs | -0.217 | 0.043 | -5.04 | <0.001 | ******* |
| Task: pd | -0.191 | 0.037 | -5.145 | <0.001 | ******* |
| Task: story | -0.472 | 0.05 | -9.36 | <0.001 | ******* |
| **P3** | **0.044** | **0.021** | **2.111** | **0.035** | ***** |
| Age | -0.009 | 0.002 | -3.918 | <0.001 | ******* |
| Sex (male) | 0 | 0.049 | -0.008 | 0.994 | — |
| Task: fs | -0.207 | 0.043 | -4.829 | <0.001 | ******* |
| Task: pd | -0.19 | 0.037 | -5.126 | <0.001 | ******* |
| Task: story | -0.471 | 0.051 | -9.327 | <0.001 | ******* |
| **N1** | **0.088** | **0.029** | **3.016** | **0.003** | ****** |
| Age | -0.023 | 0.004 | -5.56 | <0.001 | ******* |
| Sex (male) | -0.136 | 0.086 | -1.587 | 0.113 | — |
| Task: fs | -0.173 | 0.097 | -1.785 | 0.074 | — |
| Task: pd | -0.073 | 0.076 | -0.963 | 0.336 | — |
| Task: story | -0.006 | 0.09 | -0.065 | 0.948 | — |
| **N4** | **0.018** | **0.033** | **0.539** | **0.59** | — |
| Age | -0.025 | 0.004 | -5.731 | <0.001 | ******* |
| Sex (male) | -0.098 | 0.088 | -1.106 | 0.269 | — |
| Task: fs | -0.172 | 0.098 | -1.75 | 0.08 | — |
| Task: pd | -0.071 | 0.076 | -0.936 | 0.349 | — |
| Task: story | -0.005 | 0.09 | -0.052 | 0.959 | — |
| **N6** | **0** | **0.035** | **-0.011** | **0.991** | — |
| Age | -0.025 | 0.004 | -5.836 | <0.001 | ******* |
| Sex (male) | -0.092 | 0.089 | -1.036 | 0.3 | — |
| Task: fs | -0.165 | 0.098 | -1.69 | 0.091 | — |
| Task: pd | -0.071 | 0.076 | -0.932 | 0.351 | — |
| Task: story | -0.004 | 0.09 | -0.049 | 0.961 | — |
| **G5** | **0.014** | **0.057** | **0.239** | **0.811** | — |
| Age | -0.025 | 0.004 | -5.95 | <0.001 | ******* |
| Sex (male) | -0.093 | 0.088 | -1.056 | 0.291 | — |
| Task: fs | -0.169 | 0.099 | -1.715 | 0.086 | — |
| Task: pd | -0.071 | 0.076 | -0.932 | 0.351 | — |
| Task: story | -0.005 | 0.09 | -0.05 | 0.96 | — |
| **G9** | **0.047** | **0.039** | **1.201** | **0.23** | — |
| Age | -0.024 | 0.004 | -5.602 | <0.001 | ******* |
| Sex (male) | -0.097 | 0.087 | -1.112 | 0.266 | — |
| Task: fs | -0.188 | 0.099 | -1.887 | 0.059 | — |
| Task: pd | -0.07 | 0.076 | -0.919 | 0.358 | — |
| Task: story | -0.004 | 0.09 | -0.049 | 0.961 | — |
| **Total Positive** | **0.026** | **0.009** | **2.888** | **0.004** | ****** |
| Age | -0.008 | 0.002 | -3.844 | <0.001 | ******* |
| Sex (male) | -0.008 | 0.048 | -0.155 | 0.877 | — |
| Task: fs | -0.211 | 0.043 | -4.927 | <0.001 | ******* |
| Task: pd | -0.19 | 0.037 | -5.13 | <0.001 | ******* |
| Task: story | -0.472 | 0.051 | -9.342 | <0.001 | ******* |
| **Total Negative** | **0.018** | **0.012** | **1.42** | **0.156** | — |
| Age | -0.024 | 0.004 | -5.441 | <0.001 | ******* |
| Sex (male) | -0.114 | 0.088 | -1.286 | 0.199 | — |
| Task: fs | -0.178 | 0.098 | -1.823 | 0.068 | — |
| Task: pd | -0.071 | 0.076 | -0.942 | 0.346 | — |
| Task: story | -0.005 | 0.09 | -0.057 | 0.955 | — |
| **Total General** | **0.027** | **0.028** | **0.962** | **0.336** | — |
| Age | -0.024 | 0.004 | -5.598 | <0.001 | ******* |
| Sex (male) | -0.096 | 0.087 | -1.096 | 0.273 | — |
| Task: fs | -0.186 | 0.1 | -1.858 | 0.063 | — |
| Task: pd | -0.07 | 0.076 | -0.925 | 0.355 | — |
| Task: story | -0.005 | 0.09 | -0.052 | 0.959 | — |
| **PANSS Total** | **0.014** | **0.006** | **2.23** | **0.026** | ***** |
| Age | -0.022 | 0.004 | -5.12 | <0.001 | ******* |
| Sex (male) | -0.119 | 0.086 | -1.372 | 0.17 | — |
| Task: fs | -0.201 | 0.099 | -2.028 | 0.043 | * |
| Task: pd | -0.071 | 0.076 | -0.937 | 0.349 | — |
| Task: story | -0.005 | 0.09 | -0.059 | 0.953 | — |

| **Acoustic Index** |  |  |  |  |  |
| --- | --- | --- | --- | --- | --- |
| **Term** | **β** | **SE** | **z** | **p** | **Sig.** |
| **P1** | -0.035 | 0.04 | -0.858 | 0.391 | — |
| Sex (male) | -0.629 | 0.093 | -6.796 | <0.001 | ******* |
| Age | 0.015 | 0.004 | 3.621 | <0.001 | ******* |
| Task: fs | -0.041 | 0.021 | -1.948 | 0.051 | . |
| Task: pd | -0.036 | 0.018 | -1.993 | 0.046 | ***** |
| Task: story | -0.042 | 0.023 | -1.807 | 0.071 | — |
| **P2** | -0.122 | 0.042 | -2.872 | 0.004 | ****** |
| Sex (male) | -0.608 | 0.091 | -6.657 | <0.001 | ******* |
| Age | 0.013 | 0.004 | 3.196 | 0.001 | ****** |
| Task: fs | -0.04 | 0.021 | -1.905 | 0.057 | — |
| Task: pd | -0.036 | 0.018 | -1.999 | 0.046 | ***** |
| Task: story | -0.043 | 0.023 | -1.819 | 0.069 | — |
| **P3** | 0.008 | 0.04 | 0.193 | 0.847 | — |
| Sex (male) | -0.637 | 0.093 | -6.86 | <0.001 | ******* |
| Age | 0.015 | 0.004 | 3.667 | <0.001 | ******* |
| Task: fs | -0.041 | 0.021 | -1.953 | 0.051 | . |
| Task: pd | -0.036 | 0.018 | -1.99 | 0.047 | ***** |
| Task: story | -0.042 | 0.023 | -1.802 | 0.072 | — |
| **N1** | -0.091 | 0.035 | -2.557 | 0.011 | ***** |
| Sex (male) | -0.344 | 0.104 | -3.319 | <0.001 | ******* |
| Age | 0.032 | 0.005 | 6.806 | <0.001 | ******* |
| Task: fs | -0.109 | 0.051 | -2.145 | 0.032 | ***** |
| Task: pd | 0.033 | 0.04 | 0.836 | 0.403 | — |
| Task: story | 0.044 | 0.046 | 0.966 | 0.334 | — |
| **N4** | -0.095 | 0.038 | -2.492 | 0.013 | ***** |
| Sex (male) | -0.363 | 0.103 | -3.528 | <0.001 | ******* |
| Age | 0.031 | 0.005 | 6.335 | <0.001 | ******* |
| Task: fs | -0.103 | 0.051 | -2.016 | 0.044 | ***** |
| Task: pd | 0.033 | 0.04 | 0.84 | 0.401 | — |
| Task: story | 0.045 | 0.046 | 0.983 | 0.326 | — |
| **N6** | -0.067 | 0.041 | -1.645 | 0.1 | — |
| Sex (male) | -0.362 | 0.105 | -3.446 | <0.001 | ******* |
| Age | 0.032 | 0.005 | 6.457 | <0.001 | ******* |
| Task: fs | -0.105 | 0.051 | -2.06 | 0.039 | ***** |
| Task: pd | 0.033 | 0.04 | 0.821 | 0.412 | — |
| Task: story | 0.045 | 0.046 | 0.969 | 0.332 | — |
| **G5** | -0.149 | 0.066 | -2.249 | 0.025 | ***** |
| Sex (male) | -0.386 | 0.103 | -3.752 | <0.001 | ******* |
| Age | 0.032 | 0.005 | 6.687 | <0.001 | ******* |
| Task: fs | -0.101 | 0.051 | -1.978 | 0.048 | ***** |
| Task: pd | 0.033 | 0.04 | 0.824 | 0.41 | — |
| Task: story | 0.045 | 0.046 | 0.976 | 0.329 | — |
| **G9** | -0.142 | 0.045 | -3.139 | 0.002 | ****** |
| Sex (male) | -0.38 | 0.101 | -3.764 | <0.001 | ******* |
| Age | 0.03 | 0.005 | 6.193 | <0.001 | ******* |
| Task: fs | -0.097 | 0.051 | -1.908 | 0.056 | — |
| Task: pd | 0.032 | 0.04 | 0.806 | 0.421 | — |
| Task: story | 0.044 | 0.046 | 0.963 | 0.335 | — |
| **Total Positive** | -0.025 | 0.017 | -1.433 | 0.152 | — |
| Sex (male) | -0.62 | 0.093 | -6.689 | <0.001 | ******* |
| Age | 0.014 | 0.004 | 3.345 | <0.001 | ******* |
| Task: fs | -0.041 | 0.021 | -1.937 | 0.053 | . |
| Task: pd | -0.036 | 0.018 | -1.998 | 0.046 | ***** |
| Task: story | -0.043 | 0.023 | -1.815 | 0.07 | — |
| **Total Negative** | -0.038 | 0.015 | -2.625 | 0.009 | ****** |
| Sex (male) | -0.344 | 0.103 | -3.332 | <0.001 | ******* |
| Age | 0.031 | 0.005 | 6.235 | <0.001 | ******* |
| Task: fs | -0.104 | 0.051 | -2.05 | 0.04 | ***** |
| Task: pd | 0.033 | 0.04 | 0.835 | 0.404 | — |
| Task: story | 0.045 | 0.046 | 0.975 | 0.329 | — |
| **Total General** | -0.104 | 0.031 | -3.325 | <0.001 | ******* |
| Sex (male) | -0.381 | 0.1 | -3.792 | <0.001 | ******* |
| Age | 0.029 | 0.005 | 6.022 | <0.001 | ******* |
| Task: fs | -0.095 | 0.051 | -1.854 | 0.064 | — |
| Task: pd | 0.032 | 0.04 | 0.812 | 0.417 | — |
| Task: story | 0.045 | 0.046 | 0.972 | 0.331 | — |
| **PANSS Total** | -0.025 | 0.007 | -3.383 | <0.001 | ******* |
| Sex (male) | -0.348 | 0.101 | -3.444 | <0.001 | ******* |
| Age | 0.029 | 0.005 | 5.778 | <0.001 | ******* |
| Task: fs | -0.099 | 0.051 | -1.943 | 0.052 | . |
| Task: pd | 0.033 | 0.04 | 0.829 | 0.407 | — |
| Task: story | 0.045 | 0.046 | 0.969 | 0.332 | — |

**Note**. Mixed-effect models were fitted with the task-level index as the dependent variable and PANSS score as the main predictor, controlling for age, sex, and task (reference level = dream telling). Tasks: fs = free speech; pd = picture description; story = cartoon storyboard. Models were estimated using mixed-effects beta regression with a logit link and participant-specific random intercepts. Because beta regression requires values to lie strictly between 0 and 1, two observations with index values equal to 0 or 1 were excluded from the analyses. For the semantic index models, the sample size was N = 1513 observations across 221 participants for P-scale models, and N = 748 observations across 113 participants for N-scale, G-scale, and total PANSS models. For the acoustic index models, the sample size was N = 1486 observations across 221 participants for P-scale models, and N = 722 observations across 113 participants for N-scale, G-scale, and total PANSS models.  *** p < .001, ** p < .01, * p < .05.
